# Associations of adiposity, kidney stone disease, and serum calcium concentrations; observational and genetic epidemiological studies

**DOI:** 10.1101/2022.06.10.22276271

**Authors:** Catherine E Lovegrove, Jelena Bešević, Akira Wiberg, Ben Lacey, Thomas J Littlejohns, Naomi E Allen, Michelle Goldsworthy, Jihye Kim, Fadil Hannan, Gary C Curhan, Ben Turney, Mark McCarthy, Anubha Mahajan, Rajesh V Thakker, Michael V Holmes, Dominic Furniss, Sarah A Howles

**Affiliations:** Nuffield Department of Surgical Sciences, University of Oxford, Oxford, UK; Nuffield Department of Population Health, University of Oxford, Oxford, UK; Nuffield Department of Orthopaedics, Rheumatology and Musculoskeletal Sciences, University of Oxford, Oxford, UK; Department of Population Health, University of Oxford, Oxford, UK; Academic Endocrine Unit, Radcliffe Department of Medicine, University of Oxford, Oxford, UK; Harvard T.H. Chan School of Public Health, Boston, MA, USA; Nuffield Department of Women’s and Reproductive Health, University of Oxford, Oxford, UK; Channing Division of Network Medicine and Renal Division, Brigham and Women’s Hospital, Harvard Medical School, Boston, MA, USA; Wellcome Centre for Human Genetics, Nuffield Department of Medicine, University of Oxford, Oxford, UK; Medical Research Council, Integrative Epidemiology Unit, University of Bristol, Bristol, UK

**Author notes:** Corresponding author Correspondence to Sarah A. Howles.

## Abstract

**Background:** Kidney stone disease (KSD) is linked to obesity, metabolic syndrome and biochemical alterations including higher serum calcium concentration. The mechanisms by which these phenotypes associate with KSD are uncertain. We aimed to establish the effects of adiposity on KSD using conventional and genetic epidemiological techniques.

**Methods:** We assessed observational associations between measures of adiposity and incident KSD in 479,405 people from the UK Biobank. To facilitate Mendelian randomization (MR) analyses, we undertook genome-wide association studies (GWAS) of KSD in the UK Biobank in combined and sex-specific subsets. Univariable, multivariable and mediation MR analyses were used to calculate odds ratio (OR) or beta coefficient (ß) for risk of KSD per genetically instrumented higher marker of adiposity, metabolic syndrome parameter, biochemical phenotype, and inflammation and identify violations of MR assumptions.

**Findings:** Observational analyses demonstrated that measures of central adiposity (waist-to-hip ratio (WHR) and waist circumference (WC)) are more strongly associated with incident KSD than measures of general adiposity (body mass index (BMI)). Three novel KSD-GWAS loci were identified (*SLC2A12, TRPV5*, and *SLC28A1*); no sex-specific loci were detected. MR analyses established that higher central adiposity is causally linked to both KSD and higher adjusted serum calcium concentrations independent of BMI (one standard deviation higher WHR: OR for KSD=1·43, p=4·1×10^−6^; ß for serum calcium concentration=0·11mmol/L, p=2·7×10^−7^). Mediation analyses indicated that 12% of the effect of WHR on KSD is due its role in elevating serum calcium concentration. Our MR studies indicated that other components of the metabolic syndrome, serum uric acid levels, and biomarkers of inflammation are unlikely to be implicated in the causation of KSD.

**Interpretation:** Our study indicates that visceral adipose depots elevate serum calcium concentration and cause an increased risk of KSD. Therapies targeting central adipose deposition may affect calcium homeostasis and have utility for the prevention of KSD.

## Introduction

Kidney stone disease (KSD) affects ∼20% of men and ∼10% of women by 70 years of age, placing a considerable burden on health care systems and having a substantial impact on quality of life.^1,2^ Our understanding of the pathophysiology of KSD is incomplete, preventing effective prophylaxis in many cases.^2^

Observational studies indicate that both general (body mass index (BMI)) and central adiposity (waist-to-hip ratio (WHR) and waist circumference (WC)) are associated with an increased risk of KSD.^3^ Metabolic syndrome represents a cluster of conditions including central obesity, hypertension, dyslipidaemia, and impaired glucose tolerance, all of which have been linked to an increased risk of KSD.^4,5^ However, the mechanisms by which obesity and metabolic syndrome result in an increased risk of KSD remain uncertain. Pathways linking increasing adiposity with KSD may include hyperuricosuria and hyperoxaluria due to increased BMI; hyperinsulinaemia resulting in hypercalciuria; insulin resistance causing impaired renal ammonium metabolism and hypocitraturia; hypertension impacting on the urinary lithogenic profile; and vascular insult resulting in alterations in renal papillary circulation.^3,6,7^ Increasing adiposity has also been linked to alterations in serum concentrations of calcium, phosphate, vitamin D, and urate, all of which may impact on risk of KSD.^8,9^ Furthermore, it is well documented that obesity increases serum markers of systemic inflammation and inflammation is postulated to increase risk of KSD.^10^

Conventional epidemiological studies may be subject to bias, particularly from reverse causality and confounding.^11^ Mendelian randomization (MR) overcomes these problems, using genetic variants associated with an exposure to enable an unbiased estimation of causal effects and delineation of the direction of causality.^11^ Furthermore, multivariable and mediated MR facilitate identification of independent causal effects with respect to multiple exposures and estimation of relative importance.^11^ In this study, we leveraged conventional and genetic epidemiological techniques to investigate independent associations of markers of adiposity, metabolic syndrome and inflammation with KSD.

## Methods

### Study participants

The UK Biobank is a prospective study of 502,000 individuals aged 40-69 years, recruited between 2006-10, who provided informed consent, completed health-related questionnaires, provided physical measurements and blood samples, and consented to linkage of data to longitudinally collected medical records.^12^ UK Biobank is approved by the National Information Governance Board for Health and Social Care, and the National Health Service North West Centre for Research Ethics Committee (Ref:11/NW/0382). Incident KSD cases and cases for genome-wide association studies (GWAS) were identified using ICD-9, ICD-10, OPCS codes from hospital inpatient records and a self-reported code for kidney stone surgery. Prevalent cases were identified using the same codes with the addition of a self-reported condition code (Supplementary Tables 1 and 2).

### Observational analyses

Analyses excluded participants with missing or extreme (top/bottom 0.001%) values of anthropometric measurements, prevalent KSD or conditions predisposing to KSD (Supplementary Table 3). Participants were censored at earliest diagnosis of KSD, death, loss to follow up or 28/02/2018 (participants from Wales) or 31/03/2021 (participants from England and Scotland).

Cox proportional hazards regression models were used to estimate hazard ratios (HR) for associations of measures of general (BMI) and central (WHR and WC) adiposity, with incident KSD. Models were stratified by age-at-risk (in 5-year groups) and ethnicity (white, other ethnic groups), and adjusted for Townsend Deprivation Index, smoking (never, former, current), alcohol drinking (never, former, occasional, at least weekly), and, where appropriate, sex. Associations were corrected for regression dilution using the correlation between re-survey and baseline measurements, to give the associations with long-term average (“usual”) levels; or, in figures, by plotting HRs against the mean of the adiposity measure at resurvey within each baseline-defined group.^13^ Associations are therefore described between usual BMI, WHR, and WC, and incident KSD.

In categorical analyses, anthropometric measures were classified, separately in men and women, into quintiles, with HRs reported relative to the lowest category. Confidence intervals (CIs) in these analyses were calculated using variance of log risk.^14^ Linear associations are reported per 5kg/m^2^ higher usual BMI, per 0·05 higher usual WHR, and per 10cm higher usual WC. These measures were used in preference to standard deviation (SD) to facilitate comparison of estimates between male and female subsets and enable application across populations. The relative independence of different anthropometric measures were assessed by adjusting the association with BMI by measures of central adiposity (WHR and WC), and the associations with central adiposity measures by BMI. χ^2^ values were derived from likelihood ratio statistics.^15^ All analyses were performed using Rv4·1·1.

### Genome-wide association studies

Analyses excluded participants with conditions predisposing to KSD (Supplementary Table 3). Genotyping was undertaken using UK-BiLEVE and UK-Biobank Axiom Arrays which share ∼95% of marker content. Genotypes were called using array intensity data and a custom genotype-calling pipeline^16^

PLINKv1·9 and Rv3·6·1 were used for quality control. Single nucleotide polymorphisms (SNPs) with a call rate <90% were omitted. Sample-level quality control excluded individuals with one or more of: call rate <98%; discrepancy between genetic-sex (data-field 22001) and self-reported sex (data-field 21); sex chromosome aneuploidy (data-field 22019); heterozygosity >3SD from the mean (calculated using UKB PCA-adjusted heterozygosity values, data-field 20004). Individuals not of white British ancestry were excluded (based on principal component analysis and self-reporting as “British” (data-field 22006)). SNPs with Hardy–Weinberg equilibrium p<10^−4^, <98% call rate, and minor allele frequency (MAF) <1% were excluded in SNP-level quality control.

UK Biobank phasing on autosomes was performed with SHAPEIT3 with the 1000 Genomes Phase 3 dataset as a reference panel. The Haplotype Reference Consortium reference panel and a merged UK10K/1000 Genomes Phase 3 panel were used in imputation. The resultant dataset comprised 92,693,895 autosomal SNPs, short indels and large structural variants.^16^

GWAS were undertaken using 547,011 autosomal genotyped and 8,397,548 imputed SNPs of MAF≥0·01 and Info Score ≥0·9, using a linear mixed non-infinitesimal model implemented in BOLT-LMMv2·3 using the hg19 reference genetic map and reference linkage disequilibrium (LD) score file for European ancestry.^17^ Genotyping platform and sex, where appropriate, were incorporated as covariates. Quantile–quantile and Manhattan plots were generated in FUMA. Conditional analyses were performed using QCTOOLv2.

### Mendelian randomisation

MR was used to estimate causal effects of adiposity, serum, plasma, and urine parameters, estimated heel bone mineral density (BMD), and metabolic syndrome phenotypes, on KSD. SNPs with independent, GWAS significant (p<5×10^−8^) associations with phenotypes of interest in individuals of European ancestry were selected as instrumental variables from relevant studies (Supplementary Table 4). Univariable, multivariable, mediation and contamination mixture MR analyses were performed using MendelianRandomization in R.^11,18^ Where MR-Egger regression intercept estimate was zero (p>0·05), inverse-variance weighted (IVW) was interpreted as estimate of best fit. Where MR-Egger intercept estimate suggested horizontal pleiotropy (i.e. associations between instrumental variables and KSD via alternative pathways; p<0·05), MR-Egger regression was interpreted as estimate of best fit.^19^ IVW p-values were adjusted for multiple testing via the false discovery rate method. Risk of bias and type 1 error due to participant overlap between study cohorts for BMI, WHR and KSD were calculated as 0·05 and 0·016, respectively, via https://sb452.shinyapps.io/overlap/.^20^

## Results

### Associations of adiposity and kidney stone disease

Among 479,405 participants included in observational analyses, mean age at baseline was 56·5 years (SD=8·1); 55% were female (Table 1). Participants were mainly of white ethnicity (95%) and from less deprived areas than the national average (mean Townsend Deprivation Index score -1·33, SD=3·1). At baseline, mean BMI, WHR and WC were 27·8kg/m^2^ (SD=4·8), 0·94 (SD=0·06), and 96·8cm (SD=11·2), respectively, among men, and 27·1kg/m^2^ (SD=5·1), 0·82 (SD=0·07), and 84·6cm (SD=12·4), respectively, among women. Self-correlations between baseline and resurvey measurements (to correct for regression dilution bias) were 0·93 for BMI, 0·67 for WHR, and 0·82 for WC in men, and 0·92 for BMI, 0·66 for WHR, and 0·83 for WC in women.

**Table 1:** Baseline characteristics of the participants in observational studies.

|  | **Men**<br>**(n=216,157)** | **Women**<br>**(n=263,248)** | **Overall**<br>**(n=479,405)** |
| --- | --- | --- | --- |
| **Demographic and lifestyle factors** |  |  |  |
| Age, mean (SD) | 56.7 (8.2) | 56.3 (8.0) | 56.5 (8.1) |
| White, n (%) | 204,727 (94.7) | 249,298 (94.7) | 454,025 (94.7) |
| Current smoker, n (%) | 26,863 (12.4) | 23,288 (8.8) | 50,151 (10.5) |
| Alcohol drinker (at least weekly), n (%) | 168,274 (77.8) | 165,048 (62.7) | 333,322 (69.5) |
| Townsend Deprivation Index, mean (SD) | -1.3 (3.1) | -1.4 (3.0) | -1.3 (3.1) |
| **Anthropometric measures, mean (SD)** |  |  |  |
| Body mass index, kg/m<sup>2</sup> | 27.8 (4.2) | 27.1 (5.1) | 27.4 (4.8) |
| Waist circumference (cm) | 96.8 (11.2) | 84.6 (12.4) | 90.1 (13.4) |
| Waist-to-hip ratio | 0.94 (0.06) | 0.82 (0.07) | 0.87 (0.09) |
| **Kidney Stone Disease status** |  |  |  |
| Case, n (%) | 3146 (1.5) | 1830 (0.7) | 4976 (1.0) |

During 5·6 million person-years of follow-up (mean 11·6 years per person), 4,976 individuals developed incident KSD. In both sexes, each adiposity measure was strongly positively associated with incident KSD, with no evidence of a threshold throughout the ranges examined (Figure 1). There was no evidence to indicate the strength of associations differed between men and women when comparing 95% CIs for each parameter. In analyses combining both sexes, 5kg/m^2^ higher usual BMI was associated with approximately 30% higher risk of incident KSD (HR=1·31, 95% CI=1·27-1·35), as was 0·05 higher usual WHR (HR=1·34, 95% CI=1·30-1·38), and 10cm larger WC (HR=1·29, 95% CI=1·26-1·32) (Figure 2).

**Figure 1:**
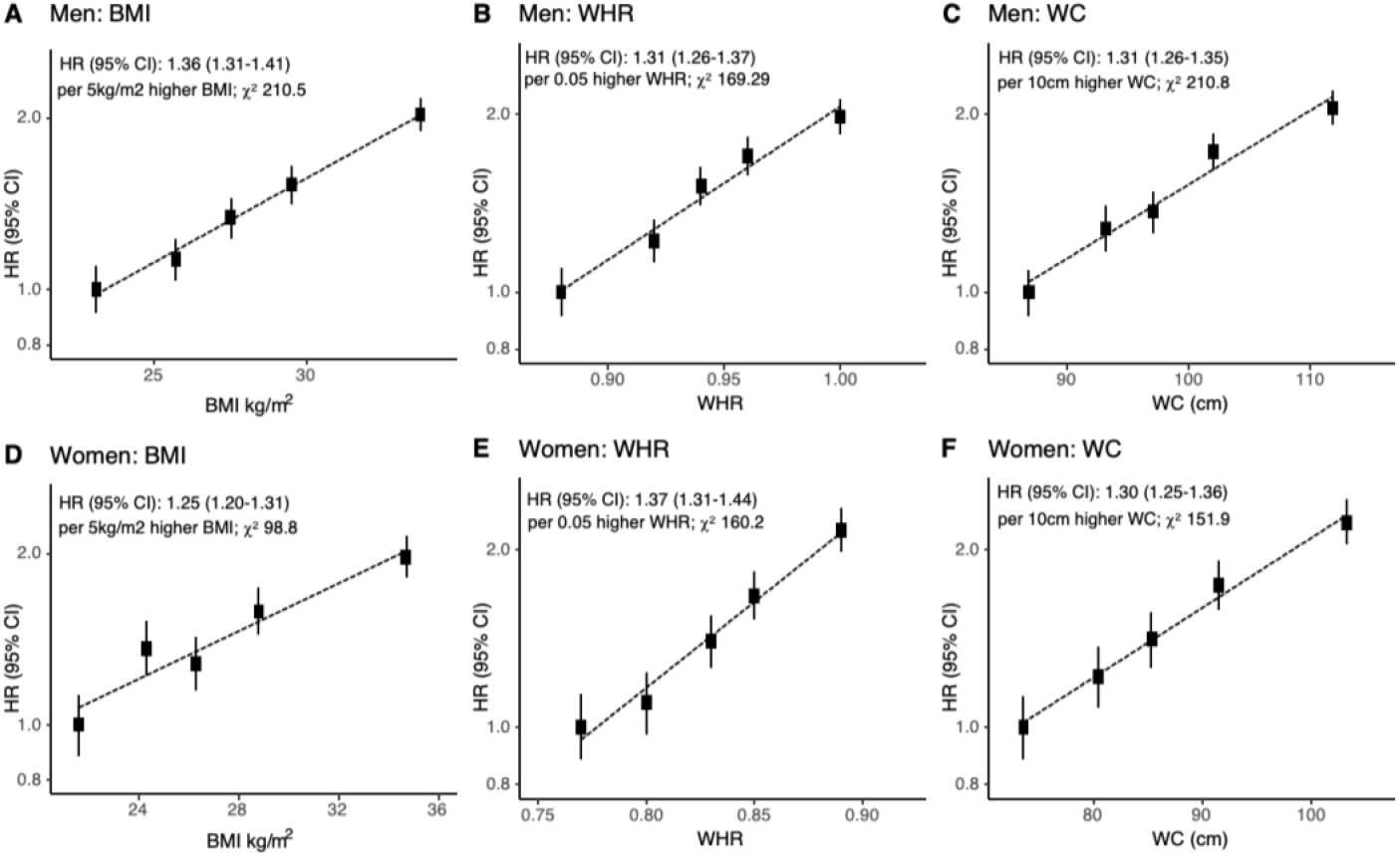
Incident kidney stone disease vs body mass index (BMI), waist-to-hip ratio (WHR) and waist circumference (WC), by sex. Hazard ratios (HR) stratified by age-at-risk and ethnicity, and adjusted for Townsend Deprivation Index, smoking and alcohol drinking, among 479,405 participants. Analyses exclude participants with pre-existing kidney stones (or conditions known to predispose to kidney stones) at baseline, and those with missing or outlying values in anthropometric variables or key covariates. The variance of the category−specific log risk determines the confidence interval (CI).

**Figure 2:**
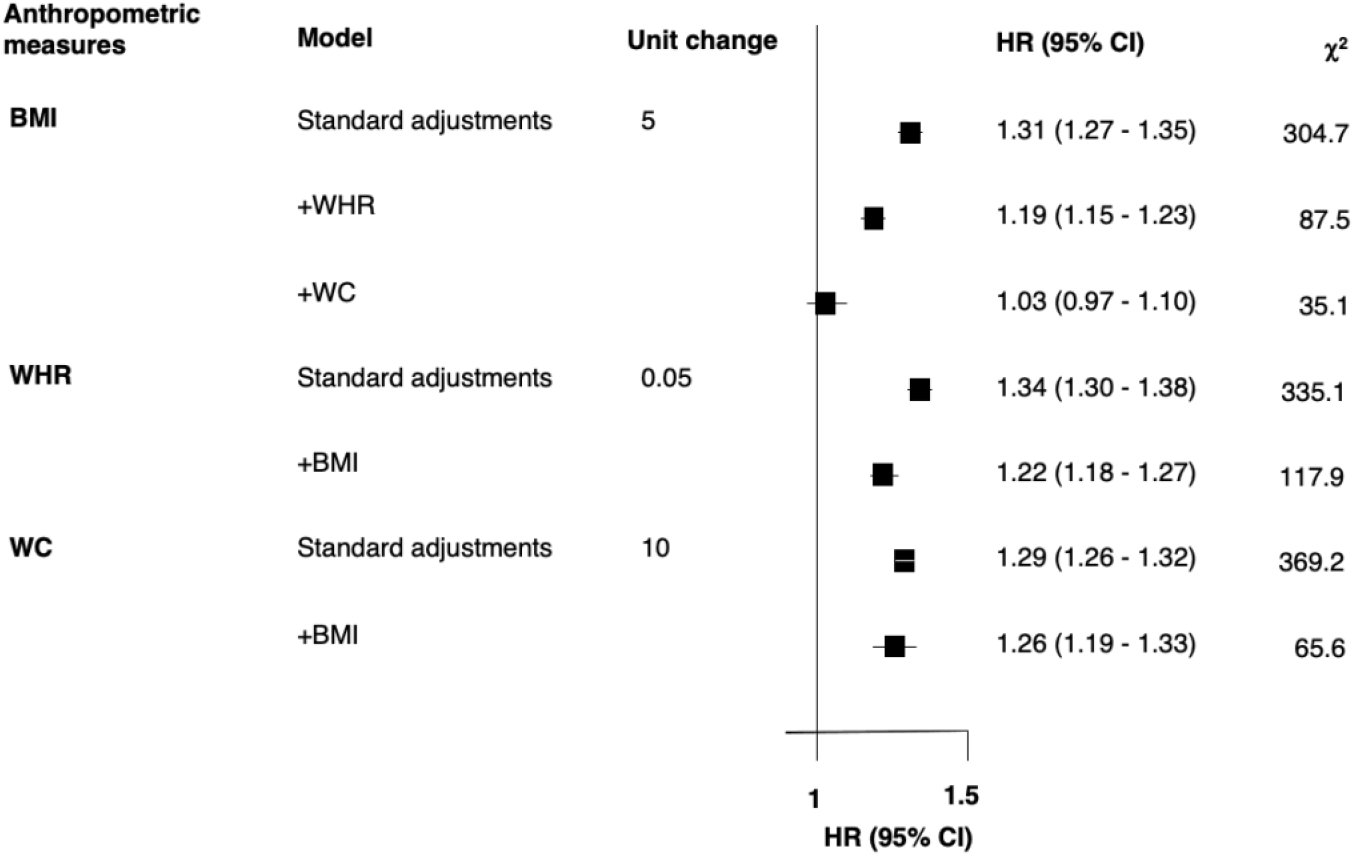
Comparison of associations between incident kidney stone disease and body mass index (BMI), waist-to-hip ratio (WHR) and waist circumference (WC), with additional adjustment for each other. Hazard ratios (HR) for the given unit change (BMI-kg/m^2^, WC-cm), stratified by age-at-risk and ethnicity, and adjusted for Townsend Deprivation Index, smoking and alcohol drinking (standard adjustments), with further adjustments for other anthropometric measures, where indicated. Analyses exclude participants with pre-existing kidney stones (or conditions known to predispose to kidney stones) at baseline, and those with missing or outlying values in anthropometric variables or key covariates, leaving 479,405 participants. The variance of the category−specific log risk determines the confidence interval (CI).

Measures of central adiposity (WHR and WC) were more strongly related to incident KSD than measures of general adiposity (as indicated by HR per one SD higher usual level of each measure; Supplementary Figure 1). In analyses of the independent effects of each adiposity measure, the association of BMI with incident KSD was almost completely attenuated following adjustment for WC (HR=1·03, 95% CI=0·97-1·10 per 5kg/m^2^), whereas both WC and WHR remained positively associated following adjustment for BMI (HR=1·26 95% CI=1·19-1·33 per 10cm higher WC, and HR=1·22, 95% CI=1·18-1·27 per 0.05 higher WHR; Figure 2).

### Genome-wide association study of kidney stone disease

To facilitate MR, we extended our previous GWAS of KSD in the UK Biobank and considered male- and female-sex specific populations including data from 8,504 participants with prevalent KSD and 388,819 controls (Supplementary Table 5).^2^

In combined-sex GWAS, 21 independent signals at 15 genetic loci associated with KSD (Figure 3, Table 2). Three novel loci were identified (*SLC2A12, TRPV5*, and *SLC28A1*). We detected a 21 base-pair in-frame deletion in *SLC34A1* previously reported to associate with KSD (OR=1.32, 95% CI=1·20-1·46, p=2·3×10^−8^).^21^ This variant results in the loss of 7 amino acids in the renally-expressed sodium-phosphate transport protein 2A (NaPi-IIa, p.Val91_Ala97del) and causes impaired membrane localisation *in vitro* (Figure 3, Table 2).^,22^ We identified that 7% of participants with KSD in the UK Biobank carry this variant compared to 5% of controls, and that it is associated with increased albumin-adjusted serum calcium (β=0·05mmol/L, 95% CI=0·03-0·07, p=2·3×10^−10^) and decreased serum phosphate (β=-0·075mmol/L, 95% CI=-0·09--0·06, p=2·3×10^−21^) concentrations in over 300,000 UK Biobank participants (Supplementary Table 4). Sex-specific GWAS identified 3 and 15 independent signals associated with KSD in female and male participants, respectively. All signals were directionally concordant; however, only *SLC34A1, UMOD, CYP24A1*, and *CLDN14* loci reached GWAS replication significance threshold (p<5·0×10^−5^) in both sexes (Supplementary Figures 2 and 3, Supplementary Tables 6 and 7).

**Figure 3:**
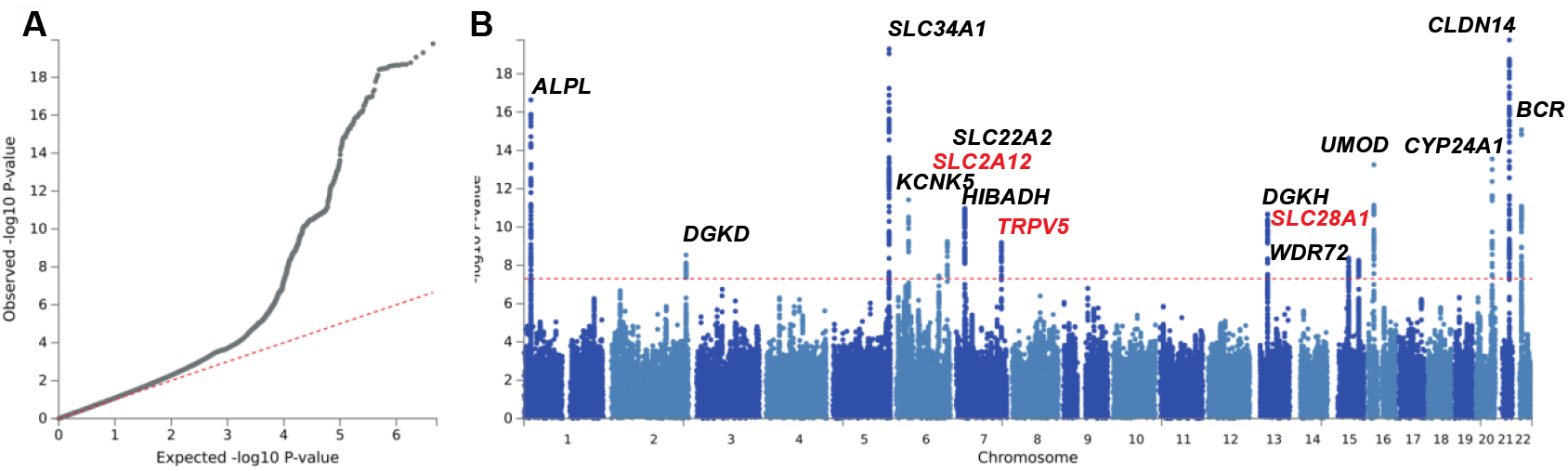
Results of combined sex genome-wide association study (GWAS) in kidney stone disease (KSD). A GWAS of KSD was performed for 8,504 individuals with KSD and 318,819 controls from the UK Biobank. **A** is a quantile-quantile plot of observed vs. expected p-values. λGC =1·001, LD score regression (LDSC) intercept= 1·01, attenuation ratio=0·31. **B** is a Manhattan plot showing the genome-wide p values (-log10) plotted against their respective positions on each of the autosomes. The horizontal red line indicates the genome-wide significance threshold of 5·0 × 10−8. Loci have been labelled with the primary candidate gene at each locus, as shown in Table 2. Previously unreported GWAS-discovered kidney stone loci are highlighted in red.

**Table 2:**
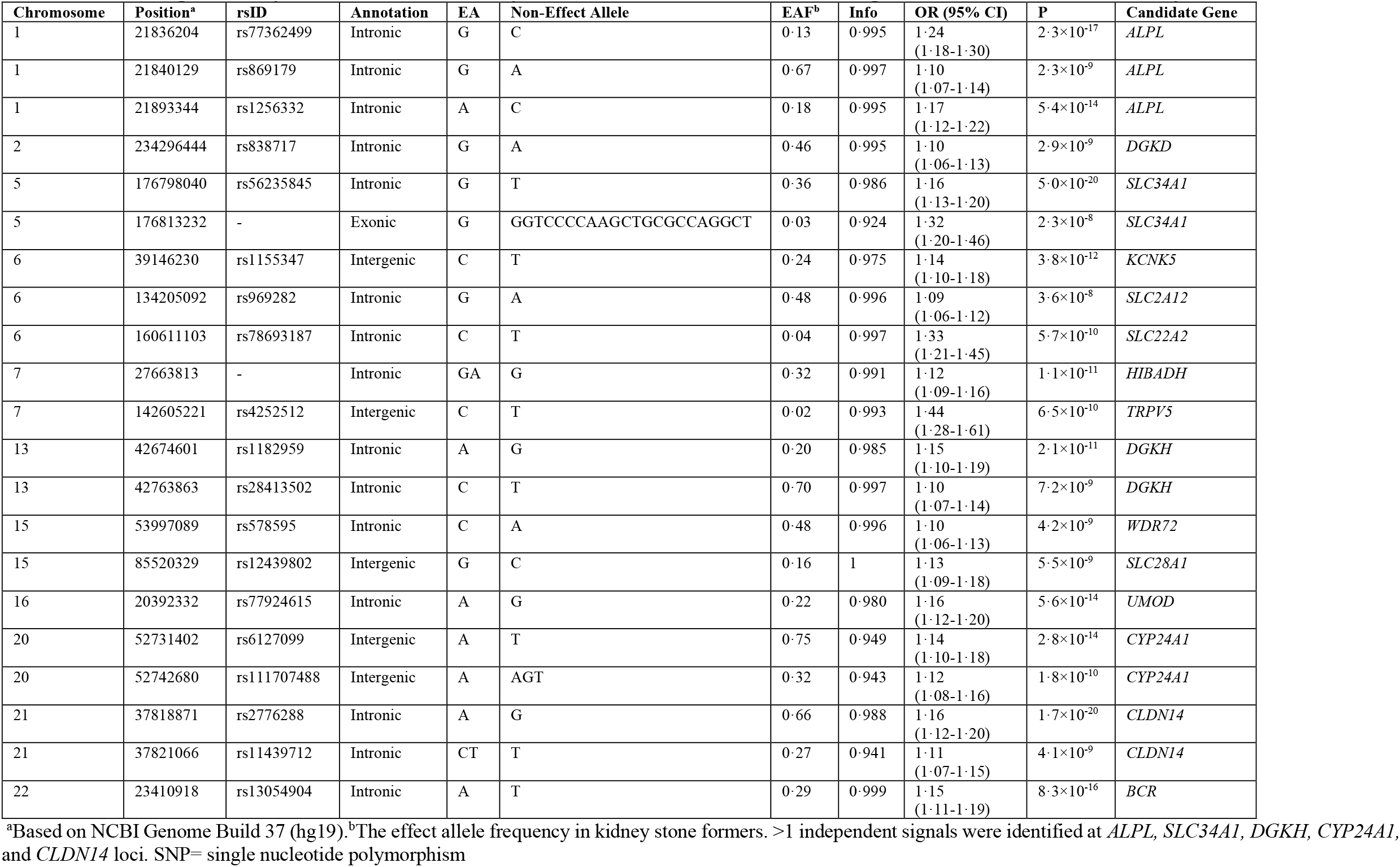
SNPs significantly associated with kidney stone disease from combined sex genome-wide association study.

### Mendelian Randomisation

In combined-sex cohorts, IVW estimates demonstrated that a one SD higher genetically instrumented BMI, WC, and WHR resulted in OR for KSD of 1·34 (95% CI=1·22-1·47, p=2·2×10^−8^), 1·47 (95% CI=1·18-1·82, p=1·6×10^−3^), and 1·46 (95% CI=1·27-1·67, p=4·1×10^−7^), respectively (Figure 4, Table 3). Multivariable MR revealed that the effect of BMI on KSD is attenuated following adjustment for WC (OR=1·08, 95% CI=0·80-1·45, p=0·71) and WHR (OR=1·20, 95% CI=1·07-1·34, p=0·01) and the effect of WC on risk of KSD is attenuated following adjustment for BMI (OR=1·37, 95% CI=0·99-1·90, p=0·13). However, higher WHR retained a positive effect on risk of KSD following adjustment for BMI (OR=1·43, 95% CI=1·24-1·64, p= 4·1×10^−6^).

**Figure 4:**
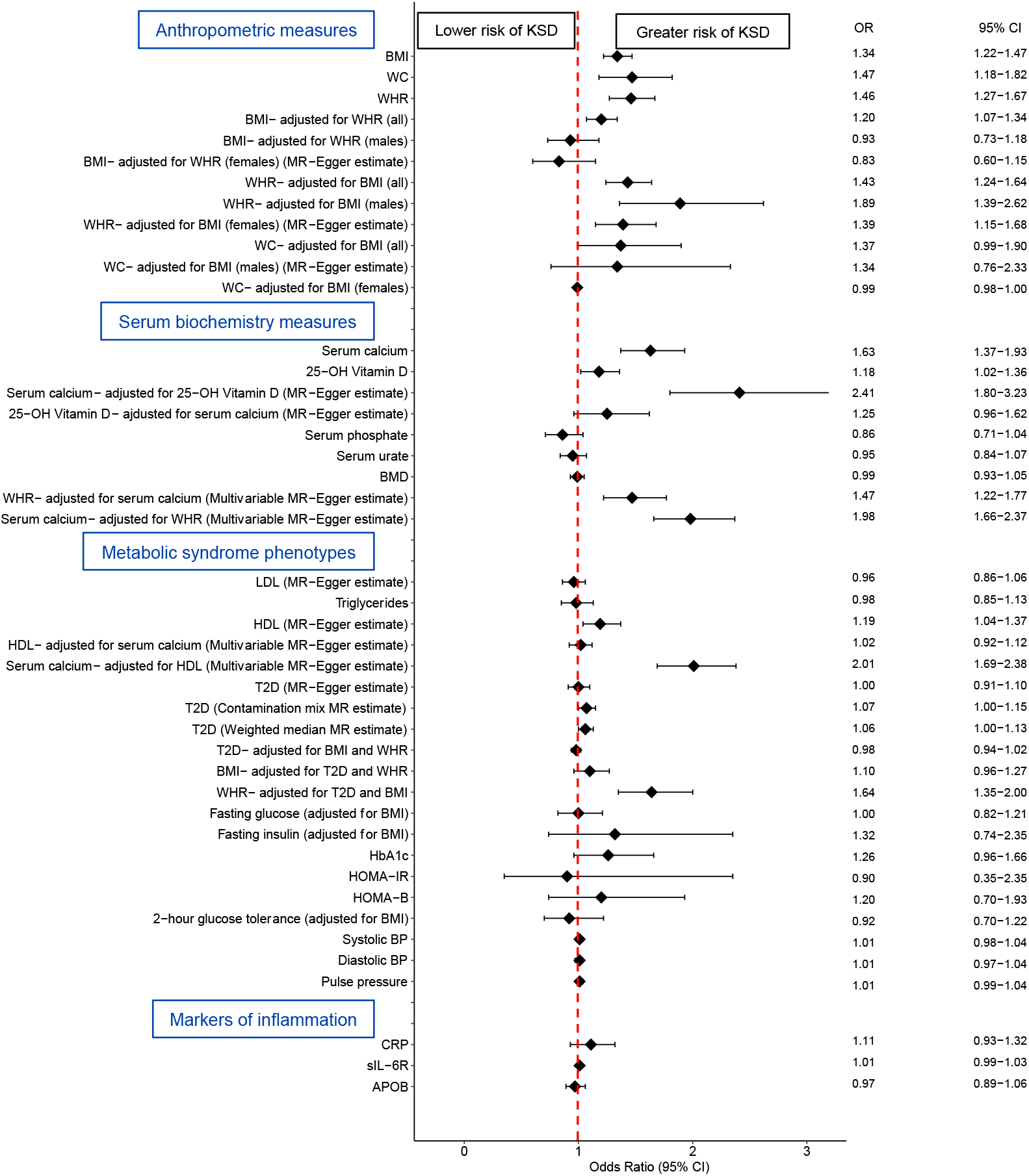
Mendelian randomization estimates for anthropomorphic, biochemical, metabolic syndrome and inflammation-related exposure variables on risk of kidney stone disease (KSD). Odds ratios (OR) and 95% confidence intervals (95% CI) of KSD per 1-standard deviation higher genetically-instrumented exposure variable. All estimates refer to inverse-variance weighted (IVW) estimates unless otherwise indicated. The vertical red line represents a line of no effect (odds ratio =1·00). All = combined male and female data, APO-B= apolipoprotein-B, BMD= bone mineral density, BMI= body mass index, CRP= C-reactive protein, HDL= high-density lipoprotein, HOMA-B= homeostasis model assessment of β-cell function, HOMA-IR= homeostasis model assessment for insulin resistance, LDL= low-density lipoprotein, sIL-6R= serum IL-6 receptor, T2D= type 2 diabetes, WC= waist circumference, WHR= waist-to-hip ratio

**Table 3:** Mendelian randomization analyses of effects of adiposity on kidney stone disease.

| Exposure | Outcome | Inverse-variance weighted |  |  | Intercept |  | MR-Egger |  |
| --- | --- | --- | --- | --- | --- | --- | --- | --- |
|  |  | OR (95% CI) | P | p-adjusted | Beta (95% CI) | P | OR (95% CI) | P |
| **BMI** | KSD (all) | **1.34**<br>(1.22-1.47) | **1.8×10<sup>-9</sup>** | **2.2×10<sup>-8</sup>** | 0.00<br>(0.00-0.00) | 0.67 | 1.40<br>(1.10-1.78) | 5.5×10<sup>-3</sup> |
|  | KSD (male) | **1.27**<br>(1.10-1.48) | **1.4×10<sup>-3</sup>** | **4.3×10<sup>-3</sup>** | 0.00<br>(-0.01-0.00) | 0.17 | 1.67<br>(1.10-2.53) | 0.02 |
|  | KSD (female) | 0.77<br>(0.64-0.92) | 5.3×10<sup>-3</sup> | 0.01 | **0.00**<br>(-0.01-0.00) | **7.5×10<sup>-3</sup>** | **1.52**<br>(0.89-2.60) | **0.13** |
| **BMI adjusted for WC (mvMR)** | KSD (all) | **1.08**<br>(0.80-1.45) | **0.62** | **0.71** | 0.00<br>(0.00-0.00) | 0.46 | **0.97**<br>(0.65-1.46) | **0.90** |
|  | KSD (male) | 1.10<br>(0.65-1.86) | 0.72 | 0.77 | **0.00**<br>(-0.01-0.00) | **0.01** | **1.70**<br>(0.92-3.17) | **0.09** |
|  | KSD (female) | 1.00<br>(0.99-1.01) | 0.64 | 0.71 | **0.00**<br>(0.00-0.00) | **3.4×10<sup>-4</sup>** | **1.02**<br>(1.00-1.03) | **8.0×10<sup>-3</sup>** |
| **BMI adjusted for WHR (mv MR)** | KSD (all) | **1.20**<br>(1.07-1.34) | **2.0×10<sup>-3</sup>** | **0.01** | 0.00<br>(0.00-0.00) | 0.48 | 1.26<br>(1.05-1.52) | 0.01 |
|  | KSD (male) | **0.93**<br>(0.73-1.18) | **0.57** | **0.71** | 0.00<br>(0.00-0.00) | 0.10 | 1.15<br>(0.82-1.61) | 0.43 |
|  | KSD (female) | 1.16<br>(0.96-1.40) | 0.13 | 0.25 | **0.00**<br>(0.00-0.00) | **0.01** | **0.83**<br>(0.60-1.15) | **0.26** |
| **WC** | KSD (all) | **1.47**<br>(1.18-1.82) | **4.6×10<sup>-4</sup>** | **1.6×10<sup>-3</sup>** | 0.00<br>(0.00-0.01) | 0.71 | 1.32<br>(0.73-2.39) | 0.36 |
|  | KSD (male) | **1.92**<br>(1.24-2.98) | **3.4×10<sup>-3</sup>** | **0.01** | 0.00<br>(-0.03-0.03) | 0.84 | 2.21<br>(0.51-9.50) | 0.29 |
|  | KSD (female) | **1.08**<br>(0.91-1.44) | **0.61** | **0.71** | 0.01<br>(0.00-0.03) | 0.12 | 0.59<br>(0.26-1.33) | 0.21 |
| **WC adjusted for BMI (mvMR)** | KSD (all) | **1.37**<br>(0.99-1.90) | **0.06** | **0.13** | 0.00<br>(0.00-0.00) | 0.46 | 1.40<br>(1.00-1.96) | 4.7×10<sup>-2</sup> |
|  | KSD (male) | 1.36<br>(0.77-2.40) | 0.28 | 0.48 | **0.00**<br>(-0.01-0.00) | **0.01** | **1.34**<br>(0.76-2.33) | **0.31** |
|  | KSD (female) | 1.00<br>(0.99-1.00) | 0.25 | 0.44 | **0.00**<br>(0.00-0.00) | **3.4×10<sup>-4</sup>** | **0.99**<br>(0.98-1.00) | **0.10** |
| **WHR** | KSD (all) | **1.46**<br>(1.27-1.67) | **5.9×10<sup>-8</sup>** | **4.1×10<sup>-7</sup>** | 0.00<br>(0.00-0.00) | 0.63 | 1.60<br>(1.08-2.37) | 0.02 |
|  | KSD (male) | **1.78**<br>(1.37-2.30) | **1.3×10<sup>-5</sup>** | **6.2×10<sup>-5</sup>** | 0.00<br>(-0.01-0.00) | 0.47 | 2.47<br>(0.97-6.27) | 0.06 |
|  | KSD (female) | **1.31**<br>(1.07-1.61) | **9.9×10<sup>-3</sup>** | **0.02** | 0.00<br>(0.00-0.01) | 0.26 | 1.01<br>(0.62-1.65) | 0.97 |
| **WHR adjusted for BMI (mvMR)** | KSD (all) | **1.43**<br>(1.24-1.63) | **7.3×10<sup>-7</sup>** | **4.1×10<sup>-6</sup>** | 0.00<br>(0.00-0.00) | 0.48 | 1.43<br>(1.24-1.65) | 6.1×10<sup>-7</sup> |
|  | KSD (male) | **1.89**<br>(1.36-2.62) | **1.3×10<sup>-4</sup>** | **5.1×10<sup>-4</sup>** | 0.00<br>(0.00-0.00) | 0.10 | 1.95<br>(1.40-2.70) | 6.5×10<sup>-5</sup> |
|  | KSD (female) | 1.37<br>(1.13-1.65) | 1.0×10<sup>-3</sup> | 3.3×10<sup>-3</sup> | **0.004**<br>(0.00-0.004) | **0.01** | **1.39**<br>(1.15-1.68) | **1.0×10<sup>-3</sup>** |
|  |  | **Beta (95% CI)** | **P** | **p-adjusted** | **Beta (95% CI)** | **P** | **Beta (95% CI)** | **P** |
| **KSD** | BMI | **-0.19**<br>(-1.04-0.64) | **0.64** | **0.71** | 0.003<br>(-0.00-0.01) | 0.42 | -1.21<br>(-3.85-1.42) | 0.36 |
| **KSD** | WHR | **0.42**<br>(-0.13-0.97) | **0.13** | **0.25** | -0.004<br>(-0.01-0.00) | 0.07 | 1.9<br>(0.23-3.58) | 0.03 |
| **KSD** | WC | **0.12**<br>(-1.4-1.83) | **0.80** | **0.84** | -0.004<br>(-0.02-0.01) | 0.43 | 2.13<br>(-2.92-7.19) | 0.41 |

All = combined male and female data; BMI = body mass index; CI=confidence interval; KSD = kidney stone disease; mvMR=multivariable Mendelian randomization; OR=odds ratio for outcome per 1 standard deviation increase in genetically-instrumented exposure variable; p-adjusted = p value adjusted for multiple testing using false discovery rate method; WC= waist circumference; WHR = waist-to-hip ratio.
Bold text highlights the sensitivity analysis to be interpreted after considering the estimate of the intercept.

Sex-specific multivariable MR analyses revealed no evidence for sex-specific effects of adiposity on risk of KSD. Thus, in males and females, higher WHR remains a significant cause of KSD following adjustment for BMI, whereas effects of BMI and WC on KSD are attenuated following adjustment (Figure 3, Table 3). No evidence was found that KSD causally increases markers of adiposity (Table 3).

These results provide evidence that higher WHR, a marker of central adiposity, is an independent causal factor in the pathogenesis of KSD. We hypothesised that this may be secondary to interplay between adiposity and alterations in serum or urinary biochemical phenotypes, features of metabolic syndrome, or inflammation, and used MR to explore these relationships.

### Effects of central adiposity on serum and urinary biochemical phenotypes

IVW estimates identified that a one SD higher genetically instrumented albumin-adjusted serum calcium (equivalent to 0·08mmol/L) led to an OR of 1·63 for KSD (95% CI=1·37-1·93, p=1·8×10^−7^). Multivariable MR demonstrated that higher WHR causally increases serum calcium concentrations independent of BMI (IVW ß=0·12mmol/L, 95% CI=0·07-0·16mmol/L, p=2·7×10^−7^). However, there was no evidence that higher BMI has a causal effect on serum calcium concentration after adjustment for WHR (IVW ß =-0·02mmol/L, 95% CI=-0·05-0·01, p=0·49) (Figure 4, Table 4).

**Table 4:**
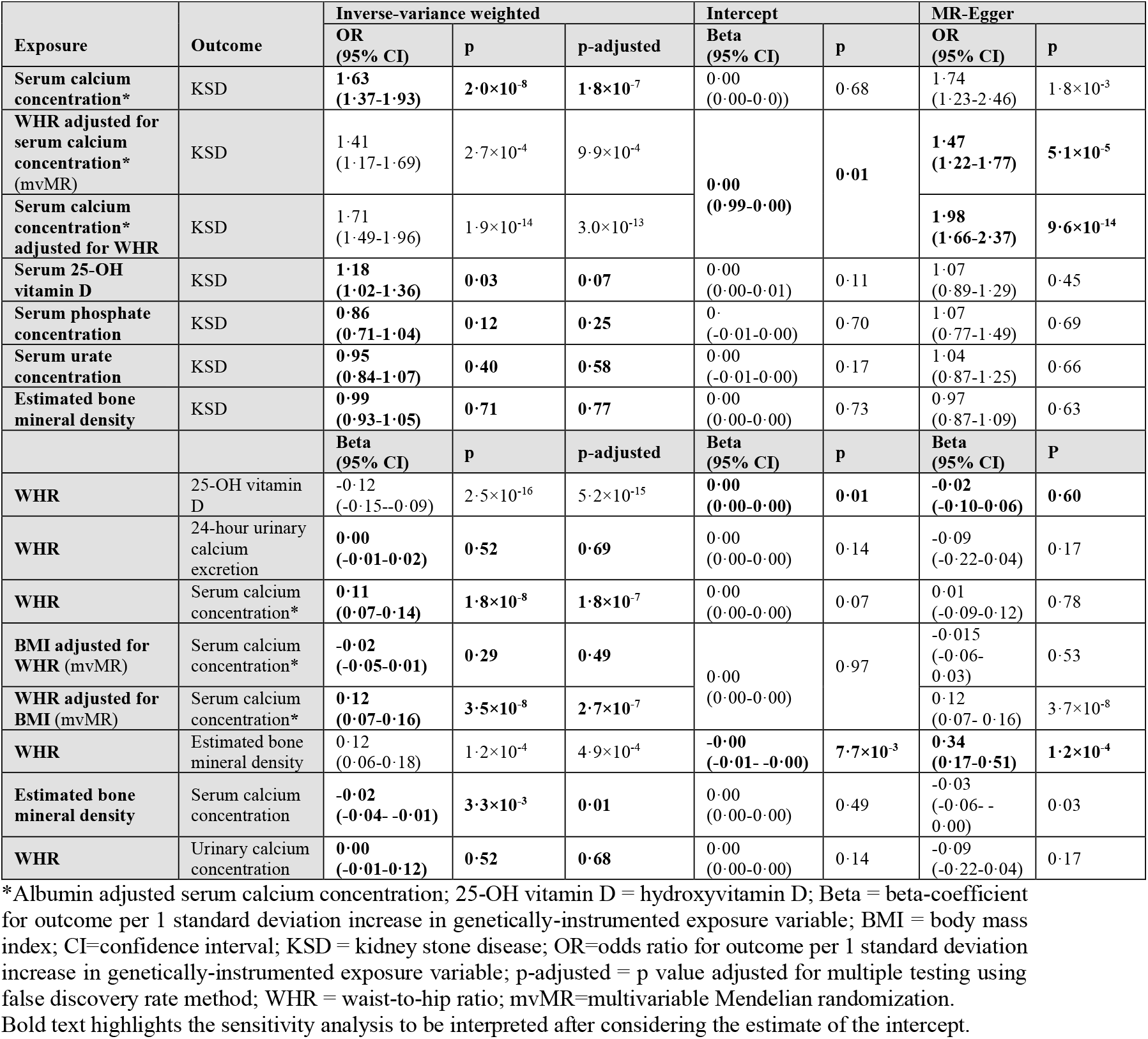
Mendelian randomization analyses of effects of serum and urinary biochemical phenotypes on kidney stone disease.

Multivariable MR indicated that higher WHR and serum calcium concentration are both independent risk factors for KSD. Thus, a one SD higher serum calcium concentration, adjusted for WHR, results in an OR of 1·98 for KSD (MR-Egger regression, 95% CI=1·10-2·37, p=9·6×10^−14^) and a one SD higher WHR, adjusted for serum calcium concentration, results in an OR of 1·47 for KSD (MR-Egger regression, 95% CI=1·22-1.77, p=5·1×10^−5^) (Figure 4, Table 4). Mediation MR suggested that 12% of the effect of WHR on KSD risk is mediated via alterations in serum calcium concentration (IVW ß=0·11mol/L, 95% CI=0·07-0·14, p=1·8×10^−8^; Supplementary Figure 4).

We hypothesized that higher WHR may alter bone resorption to cause increased serum calcium concentration. Higher WHR increases estimated heel BMD (MR-Egger ß=0·34g/cm^2^, 95% CI=0·17-0·51, p=1·2×10^−4^); however, higher BMD decreases serum calcium concentrations and does not affect risk of KSD (Figure 4, Table 4).

A one SD higher 25-OH vitamin D concentration, equivalent to 21ng/mL, may increase risk of KSD by 18%; however, after adjusting for multiple testing the p-value did not reach significance (IVW OR=1·18, 95% CI=1·02-1·36, p=0·07, Figure 4). No evidence for causal effects of higher WHR on 25-OH vitamin D concentration were detected. Neither serum phosphate nor urate demonstrated causal effects on risk of KSD. Only one independent SNP was identified from a GWAS of 24-hour urinary calcium excretion precluding confirmation via MR that higher urinary calcium excretion has a causal relationship with KSD (Supplementary Table 4). However, WHR was not found to alter urinary calcium excretion (Table 4).

These findings indicate that higher concentrations of serum calcium are causally implicated in the pathogenesis of KSD. Moreover, higher WHR results in higher serum calcium concentration, and this represents one pathway by which higher WHR may increase risk of KSD.

### Effects of components of the metabolic syndrome and markers of inflammation on kidney stone disease

No evidence was found that low-density lipoprotein (LDL) cholesterol or triglyceride serum concentrations are causally linked to KSD. However, higher high-density lipoprotein (HDL) cholesterol serum concentration was found to increase risk of KSD (MR-Egger OR=1·19, 95% CI=1·04-1·37, p=9·9×10^−3^, Figure 4, Supplementary Table 8).

A one SD higher genetically instrumented HDL causes a 0·04mmol/L increase in albumin-adjusted serum calcium concentration (95% CI=0·02-0·07mmol/L, p=2·4×10^−4^) and higher serum concentration of HDL decreases 25-OH vitamin D serum concentration (MR-Egger ß=-0·06ng/mL, 95% CI=-0·10-0·03, p=1·9×10^−4^). Multivariable MR demonstrated that following adjustment for serum calcium concentration, there is no evidence that HDL has a causal effect on the risk of KSD (Figure 4, Supplementary Table 8). These findings suggest that the effect of HDL on risk of KSD is mediated via alterations in serum calcium concentration. IVW estimates indicate that higher WHR leads to lower, rather than higher, HDL concentration (Figure 4, Supplementary Table 8). Thus, we found no evidence that increasing central adiposity causes alterations in lipid profiles to increase risk of KSD. IVW and MR-Egger analyses revealed divergent evidence regarding the causal relationship between genetic liability to type 2 diabetes (T2D) and KSD (Supplementary Table 8). Previous studies in UK Biobank and FinnGen have reported positive causal effects of T2D on risk of KSD.^23^ We therefore extended our analyses to include contamination mixture and weighted median MR approaches, identifying a potential causal effect of T2D on KSD (weighted median OR=1·06, 95% CI=1·00-1·13, p=0·06, contamination mixture OR=1·06, 95% CI=1·00-1·15, p=0·06, Supplementary Table 8). To account for potential bias from overlap of BMI-associated variants with T2D genetic instruments, we undertook multivariable MR adjusting for BMI, WHR, and T2D simultaneously. These results indicate that whilst there is some evidence that genetic liability to T2D may increase risk of KSD, this is likely confounded by co-existing adiposity (T2D following adjustment for BMI and WHR, IVW OR=0·98, 95% CI=0·94-1·02, p=0·52; BMI following adjustment for T2D and WHR IVW OR=1·10, 95% CI=0·96-1·27, p=0·36; WHR following adjustment for T2D and BMI IVW OR=1·64, 95% CI=1·35-2·00, p=3·1×10^−6^; Figure 4, Supplementary Table 8). Furthermore, we found no evidence to support causal effects of the phenotypic facets of T2D, including fasting glucose, fasting insulin, HbA1c, homeostasis model assessment of insulin resistance (HOMA-IR), homeostasis model assessment of ß-cell function (HOMA-B), and 2-hour glucose tolerance, on KSD (Figure 4, Supplementary Table 8).

No evidence was found for causal effects of hypertension, higher serum concentrations of C-reactive protein, IL-6 receptor, or apolipoprotein B on KSD (Figure 4, Supplementary Tables 8 and 9).

## Discussion

This study uses conventional and genetic epidemiological techniques to demonstrate that both central and general adiposity are risk factors for KSD, but that increasing central adiposity likely has a more important role. In both conventional and genetic analyses, we found that a one SD higher WHR results in ∼30-45% increased risk of KSD and that following adjustment for measures of central adiposity, the effects of BMI on risk of KSD were markedly attenuated. Furthermore, we demonstrate that higher WHR causes elevation of serum calcium concentrations and that a 0·08mmol/L higher serum calcium concentration causes a 63% increased risk of KSD. Using mediation MR we show that this previously unreported pathogenic mechanism mediates 12% of the effect of increasing WHR on risk of KSD risk.

Adipose tissue has multiple functions including energy storage, integration of glucose homeostasis and endocrine activity. Visceral distribution of white adipose tissue leads to increased central adiposity and is associated with metabolic disease. Our finding that central adiposity is a better discriminator of KSD risk than generalised adiposity is in keeping with studies of adipose tissue distribution and phenotypes including all-cause mortality, cardiovascular disease, and cancer-risk.^24,25^ We predict that the effects of central adiposity on calcium homeostasis and risk of KSD are related to the specific transcriptional and adipokine profiles of visceral adipose depots which may impact on PTH production; studies are required to investigate these mechanisms further and shed light on potential therapeutic targets.^26^

We found that a 21ng/mL increase in serum 25-OH vitamin D concentration may increase risk of KSD by 18%. We have previously reported that vitamin D sensitivity may be a common cause of KSD due alterations in expression of *CYP24A1*.^2^ In this study we identified an association of an in-frame deletion in *SLC34A1* with KSD, increased albumin-adjusted serum calcium and decreased serum phosphate concentrations. Biallelic loss-of-function mutations in NaPi-IIa result in urinary phosphate wasting, reduced FGF23 concentrations, activation of CYP27B1 and inhibition of CYP24A1 leading to increased serum calcium concentrations.^22^ We report that 7% of KSD UK Biobank participants carry this deletion compared to 5% of controls and predict that vitamin D supplementation may exacerbate the lithogenic profile of these individuals. Our findings indicate that genotypic assessment may be beneficial in KSD patients prior to vitamin D supplementation and that phosphate supplementation may be a useful precision-medicine approach in KSD patients carrying this *SLC34A1* deletion; further studies are required.

KSD has been described as a sexually dimorphic condition.^1^ In this study we found no evidence of genetic or anthropometric sex-specific risk factors. A recent assessment of epidemiological trends in KSD from 2007-2016 indicates the prevalence of KSD has increased in female individuals but remained static in males.^1^ This may be due to increasing rates of obesity in females, indicating that lifestyle factors, including variations in adiposity, may have driven the previously reported sex differences in KSD prevalence.^27^

It has been postulated that obesity is linked to risk of KSD due to associations with components of metabolic syndrome.^4,5^ Our study finds no evidence to support causal effects of hypertension or metabolic syndrome-associated dyslipidaemia on KSD risk. Whilst we find limited evidence that genetic liability to T2D may increase risk of KSD, we propose that these results are confounded by co-existing adiposity. These conclusions contrast with a study which demonstrated that the causal effects of adiposity on chronic kidney disease (CKD) are largely mediated by T2D, blood pressure, and their correlates; highlighting the different causal architectures of CKD and KSD.^28^ We found no evidence for causal effects of markers of systemic inflammation or serum urate concentrations on KSD.

The use of UK Biobank genetic data has enabled us to estimate causal risk factors for KSD. However, restricting these investigations to people of European ancestry may impact on applicability across populations. Furthermore, despite limited evidence of violations of MR assumptions, bias may still exist; for example, variants associated with adiposity have been shown to enrich for central nervous system cell types.^29^ It is plausible that variants included in genetic instruments may affect behaviours, such as fluid ingestion as well as energy intake, impacting on risk of KSD. In addition, some analyses may be underpowered to identify causal effects; for example, variability in 24-hour urinary excretion values may explain why higher WHR was not found to affect urinary calcium excretion.

In summary, this study reveals that higher central adiposity causes an increased risk of KSD. We found no evidence to support independent causal effects of serum urate, phosphate, T2D, hypertension, metabolic syndrome-associated dyslipidaemia, or markers of inflammation on risk of KSD. However, we demonstrate that higher central adiposity causes elevation of serum calcium concentration, and that higher serum calcium concentration causally increases risk of KSD. We predict that therapies targeting central adiposity may affect calcium homeostasis and have utility for the prevention of KSD.

## Data Availability

Individual participant data utilised in the preparation of this manuscript is available via the UK Biobank. Summary statistics for novel GWAS will be available via figshare at the time of publication.

## Acknowledgements

This research was conducted using the UK Biobank Resource under application number 885. Work was supported by grants from Kidney Research UK (RP_030_20180306) to S.A.H., A.W., M.G., B.W.T., and D.F., National Institute for Health Research (N.I.H.R) Oxford Biomedical Research Centre to R.V.T and D.F., and the Wellcome Trust to S.A.H, and M.G. (204826/z/16/z), and R.V.T. (106995/z/15/z). C.E.L. is a N.I.H.R Academic Clinical Fellow. A.W. is an N.I.H.R Academic Clinical Lecturer. S.A.H. is a Wellcome Trust Clinical Career Development Fellow. J.B., B.L. and N.E.A. acknowledge support from UK Biobank (which is a charitable company largely funded by the Medical Research Council and Wellcome Trust). M.M. and A.M. are employees of Genetech and hold stock in Roche. M.V.H. was supported by a British Heart Foundation Intermediate Clinical Research Fellowship (FS/18/23/33512) when involved in this work. M.V.H. is an employee of 23andMe and holds stock in 23andMe. G.C. acknowledges support from the National Institutes of Health (R01 DK115727 and K24 DK091417) and is an employee of OM1, Inc.

## Author Contributions

C.E.L., J.B., A.W., B.L., T.L., N.A., A.M., F.H., B.T., M.M., R.V.T., M.H, D.F., and S.A.H designed this study. C.E.L., J.B., B.L., T.L., N.A., A.W., M.G., J.K., F.H., G.C., A.M., M.H.,

D.F., and S.A.H. analyzed and interpreted data. C.E.L., J.B., and S.A.H. wrote the first draft of the manuscript. All other co-authors participated in the preparation of the manuscript by reading and commenting on the draft prior to submission.

## References

1. Abufaraj M, Xu T, Cao C, et al. Prevalence and Trends in Kidney Stone Among Adults in the USA: Analyses of National Health and Nutrition Examination Survey 2007-2018 Data. Eur Urol Focus 2021; 7(6): 1468–75.

2. Howles SA, Wiberg A, Goldsworthy M, et al. Genetic variants of calcium and vitamin D metabolism in kidney stone disease. Nat Commun 2019; 10(1): 5175.

3. Taylor EN, Stampfer MJ, Curhan GC. Obesity, weight gain, and the risk of kidney stones. JAMA 2005; 293(4): 455–62.

4. Ping H, Lu N, Wang M, et al. New-onset metabolic risk factors and the incidence of kidney stones: a prospective cohort study. BJU international 2019; 124(6): 1028–33.

5. Masterson JH, Woo JR, Chang DC, et al. Dyslipidemia is associated with an increased risk of nephrolithiasis. Urolithiasis 2015; 43(1): 49–53.

6. Borghi L, Meschi T, Guerra A, et al. Essential arterial hypertension and stone disease. Kidney international 1999; 55(6): 2397–406.

7. Stoller ML, Meng MV, Abrahams HM, Kane JP. The primary stone event: a new hypothesis involving a vascular etiology. J Urol 2004; 171(5): 1920–4.

8. Jafari-Giv Z, Avan A, Hamidi F, et al. Association of body mass index with serum calcium and phosphate levels. Diabetes & metabolic syndrome 2019; 13(2): 975–80.

9. Dalbeth N, Allan J, Gamble GD, et al. Effect of body mass index on serum urate and renal uric acid handling responses to an oral inosine load: experimental intervention study in healthy volunteers. Arthritis Res Ther 2020; 22(1): 259.

10. Amin R, Asplin J, Jung D, et al. Reduced active transcellular intestinal oxalate secretion contributes to the pathogenesis of obesity-associated hyperoxaluria. Kidney international 2018; 93(5): 1098–107.

11. Carter AR, Sanderson E, Hammerton G, et al. Mendelian randomisation for mediation analysis: current methods and challenges for implementation. European journal of epidemiology 2021; 36(5): 465–78.

12. Sudlow C, Gallacher J, Allen N, et al. UK biobank: an open access resource for identifying the causes of a wide range of complex diseases of middle and old age. PLoS medicine 2015; 12(3): e1001779.

13. Clarke R, Shipley M, Lewington S, et al. Underestimation of risk associations due to regression dilution in long-term follow-up of prospective studies. American journal of epidemiology 1999; 150(4): 341–53.

14. Easton DF, Peto J, Babiker AG. Floating absolute risk: an alternative to relative risk in survival and case-control analysis avoiding an arbitrary reference group. Stat Med 1991; 10(7): 1025–35.

15. Parish S, Peto R, Palmer A, et al. The joint effects of apolipoprotein B, apolipoprotein A1, LDL cholesterol, and HDL cholesterol on risk: 3510 cases of acute myocardial infarction and 9805 controls. European heart journal 2009; 30(17): 2137–46.

16. Bycroft C, Freeman C, Petkova D, et al. The UK Biobank resource with deep phenotyping and genomic data. Nature 2018; 562(7726): 203–9.

17. Loh PR, Kichaev G, Gazal S, Schoech AP, Price AL. Mixed-model association for biobank-scale datasets. Nature genetics 2018; 50(7): 906–8.

18. Yavorska OO, Burgess S. MendelianRandomization: an R package for performing Mendelian randomization analyses using summarized data. International journal of epidemiology 2017; 46(6): 1734–9.

19. Bowden J, Del Greco MF, Minelli C, Davey Smith G, Sheehan N, Thompson J. A framework for the investigation of pleiotropy in two-sample summary data Mendelian randomization. Stat Med 2017; 36(11): 1783–802.

20. Burgess S, Davies NM, Thompson SG. Bias due to participant overlap in two-sample Mendelian randomization. Genetic epidemiology 2016; 40(7): 597–608.

21. Sun BB, Kurki MI, Foley CN, et al. Genetic associations of protein-coding variants in human disease. Nature 2022; 603(7899): 95–102.

22. Schlingmann KP, Ruminska J, Kaufmann M, et al. Autosomal-Recessive Mutations in SLC34A1 Encoding Sodium-Phosphate Cotransporter 2A Cause Idiopathic Infantile Hypercalcemia. Journal of the American Society of Nephrology : JASN 2016; 27(2): 604–14.

23. Yuan S, Larsson SC. Assessing causal associations of obesity and diabetes with kidney stones using Mendelian randomization analysis. Molecular genetics and metabolism 2021; 134(1-2): 212–5.

24. Jayedi A, Soltani S, Zargar MS, Khan TA, Shab-Bidar S. Central fatness and risk of all cause mortality: systematic review and dose-response meta-analysis of 72 prospective cohort studies. Bmj 2020; 370: m3324.

25. Barberio AM, Alareeki A, Viner B, et al. Central body fatness is a stronger predictor of cancer risk than overall body size. Nat Commun 2019; 10(1): 383.

26. Bolland MJ, Grey AB, Gamble GD, Reid IR. Association between primary hyperparathyroidism and increased body weight: a meta-analysis. The Journal of clinical endocrinology and metabolism 2005; 90(3): 1525–30.

27. Wang Y, Beydoun MA, Min J, Xue H, Kaminsky LA, Cheskin LJ. Has the prevalence of overweight, obesity and central obesity levelled off in the United States? Trends, patterns, disparities, and future projections for the obesity epidemic. International journal of epidemiology 2020; 49(3): 810–23.

28. Zhu P, Herrington WG, Haynes R, et al. Conventional and Genetic Evidence on the Association between Adiposity and CKD. Journal of the American Society of Nephrology : JASN 2021; 32(1): 127–37.

29. Timshel PN, Thompson JJ, Pers TH. Genetic mapping of etiologic brain cell types for obesity. Elife 2020; 9.

