## Supplemental material for "Associations of adiposity, kidney stone disease, and serum calcium concentrations; observational and genetic epidemiological studies"

**Supplementary information**
**This appendix has been provided by the authors to give additional information about their work**

**Supplement to:**
**Associations of adiposity, kidney stone disease and serum calcium concentrations; observational and genetic epidemiological studies**

Contents

**Supplementary Table 1: Inclusion criteria for observational analyses** ... 3

**Supplementary Table 2: Inclusion criteria for genome-wide association analyses** ... 4

**Supplementary Table 3: Exclusion criteria for observational and genome-wide association analyses** ... 5

**Supplementary Table 4: Data sources for exposure and outcome variables in Mendelian randomization and association analyses** ... 6

**Supplementary Table 5: UK Biobank Study Population for genome-wide association studies.** ... 8

**Supplementary Table 6: Single nucleotide polymorphisms significantly associated with kidney stone disease from male sex genome-wide association study** ... 9

**Supplementary Table 7: Single nucleotide polymorphisms significantly associated with kidney stone disease from female sex genome-wide association study** ... 10

**Supplementary Table 8: Mendelian randomization analyses of effects of the metabolic syndrome on kidney stone disease** 11

**Supplementary Figure 1: Hazard ratio (HR) for incident kidney stone disease (KSD) and 95% confidence interval (CI) per standard deviation (SD) change in body mass index (BMI), waist-to-hip ratio (WHR) and waist circumference (WC)** ... 13

**Supplementary Figure 2: Results of sex-specific genome wide association study in kidney stone disease** ... 14

**Supplementary Figure 3: Sex-specific odds ratios, 95% confidence intervals and p values for significant hits in genome wide association studies in males or females.** ... 15

**Supplementary Figure 4: Effects of waist-to-hip ratio on serum calcium concentration and kidney stone disease.** ... 16

**Supplementary Figure 5: Significant relationships identified on Mendelian randomization** ... 19

### Supplementary Table 1: Inclusion criteria for observational analyses

| **International Classification of Diseases Codes (ICD)** | |
|---|---|
| ICD9- 5920 | Calculus of kidney |
| ICD9- 5921 | Calculus of ureter |
| ICD9- 5929 | Urinary calculus, unspecified |
| ICD9- 7880 | Renal colic |
| ICD10- N20.0 | Calculus of kidney |
| ICD10- N20·1 | Calculus of ureter |
| ICD10- N20·2 | Calculus of kidney with calculus of ureter |
| ICD10- N20·9 | Urinary calculus, unspecified |
| ICD10- N23 | Unspecified Renal Colic |

| **OPCS Classification of Interventions and Procedures Codes (OPCS)** | |
|---|---|
| OPCS3- 5631 | Removal of renal calculus: nephrolithomy |
| OPCS3- 5632 | Removal of renal calculus: pyelolithomy |
| OPCS3- 5633 | Removal of renal calculus: removal without incision |
| OPCS4- M06.1 | Open removal of calculus from kidney |
| OPCS4- M09·1 | Endoscopic ultrasound fragmentation of calculus of kidney |
| OPCS4- M09·2 | Endoscopic electrohydraulic shock wave fragmentation of calculus of kidney |
| OPCS4- M09·3 | Endoscopic laser fragmentation of calculus of kidney |
| OPCS4- M09·4 | Endoscopic extraction of calculus of kidney |
| OPCS4- M09·8 | Other specified therapeutic endoscopic operations on calculus of kidney, |
| OPCS4- M09·9 | Unspecified therapeutic endoscopic operations on calculus of kidney |
| OPCS4- M14·1 | Extracorporeal shock wave lithotripsy of calculus of kidney |
| OPCS4- M14·8 | Other specified extracorporeal fragmentation of calculus of kidney |
| OPCS4- M14·9 | Unspecified extracorporeal fragmentation of calculus of kidney |
| OPCS4- M16·4 | Percutaneous nephrolithotomy |
| OPCS4- M27·1 | Ureteroscopic laser fragmentation of calculus of ureter |
| OPCS4- M27·2 | Ureteroscopic fragmentation of calculus of ureter |
| OPCS4- M27·3 | Ureteroscopic extraction of calculus of ureter |
| OPCS4- M28·1 | Endoscopic laser fragmentation of calculus of ureter |
| OPCS4- M28·2 | Endoscopic fragmentation of calculus of ureter |
| OPCS4- M28·3 | Endoscopic extraction of calculus of ureter |
| OPCS4- M28·4 | Endoscopic catheter drainage of calculus of ureter |
| OPCS4- M28·5 | Endoscopic drainage of calculus of ureter by dilation of ureter |
| OPCS4- M28·8 | Other specified other endoscopic removal of calculus from ureter |
| OPCS4- M28·9 | Unspecified other endoscopic removal of calculus from ureter |
| OPCS4- M31·1 | Extracorporeal shock wave lithotripsy of calculus of ureter |
| OPCS4- M31·8 | Other specified extracorporeal fragmentation of calculus of ureter |
| OPCS4- M31·9 | Unspecified extracorporeal fragmentation of calculus of ureter |
| OPCS4- M26·1 | Nephroscopic laser fragmentation of calculus of ureter |
| OPCS4- M26·2 | Nephroscopic fragmentation of calculus of ureter NEC |
| OPCS4-M26·3 | Nephroscopic extraction of calculus of ureter |

| **Self-reported operation codes** | |
|---|---|
| 1197 | Percutaneous/open kidney stone surgery/lithotripsy |

| **Self reported condition** | |
|---|---|
| 1197 | Kidney stone/ureter stone/bladder stone |

| **Death** | |
|---|---|
| N20·0 | Calculus of kidney |
| N20·1 | Calculus of ureter |
| N20·2 | Calculus of kidney with calculus of ureter |
| N20·9 | Urinary calculus, unspecified |
| N23 | Unspecified Renal Colic |

### Supplementary Table 2: Inclusion criteria for genome-wide association analyses

| International Classification of Diseases Codes (ICD) | | OPCS Classification of Interventions and Procedures Codes (OPCS) | | Self-reported operation code | | Death | |
|---|---|---|---|---|---|---|---|
| ICD9- 5920 | Calculus of kidney | OPCS3- 5631 | Removal of renal calculus: nephrolithomy | 1197 | Percutaneous/open kidney stone surgery/lithotripsy | N20·0 | Calculus of kidney |
| ICD9- 5921 | Calculus of ureter | OPCS3- 5632 | Removal of renal calculus: pyelolithomy | | | N20·1 | Calculus of ureter |
| ICD9- 5929 | Urinary calculus, unspecified | OPCS3- 5633 | Removal of renal calculus: removal without incision | | | N20·2 | Calculus of kidney with calculus of ureter |
| ICD9- 7880 | Renal colic | OPCS4- M06·1 | Open removal of calculus from kidney | | | N20·9 | Urinary calculus, unspecified |
| ICD10- N20·0 | Calculus of kidney | OPCS4- M09·1 | Endoscopic ultrasound fragmentation of calculus of kidney | | | N23 | Unspecified Renal Colic |
| ICD10- N20·1 | Calculus of ureter | OPCS4- M09·2 | Endoscopic electrohydraulic shock wave fragmentation of calculus of kidney | | | | |
| ICD10- N20·2 | Calculus of kidney with calculus of ureter | OPCS4- M09·3 | Endoscopic laser fragmentation of calculus of kidney | | | | |
| ICD10- N20·9 | Urinary calculus, unspecified | OPCS4- M09·4 | Endoscopic extraction of calculus of kidney | | | | |
| ICD10- N23 | Unspecified Renal Colic | OPCS4- M09·8 | Other specified therapeutic endoscopic operations on calculus of kidney, | | | | |
| | | OPCS4- M09·9 | Unspecified therapeutic endoscopic operations on calculus of kidney | | | | |
| | | OPCS4- M14·1 | Extracorporeal shock wave lithotripsy of calculus of kidney | | | | |
| | | OPCS4- M14·8 | Other specified extracorporeal fragmentation of calculus of kidney | | | | |
| | | OPCS4- M14·9 | Unspecified extracorporeal fragmentation of calculus of kidney | | | | |
| | | OPCS4- M16·4 | Percutaneous nephrolithotomy | | | | |
| | | OPCS4- M27·1 | Ureteroscopic laser fragmentation of calculus of ureter | | | | |
| | | OPCS4- M27·2 | Ureteroscopic fragmentation of calculus of ureter | | | | |
| | | OPCS4- M27·3 | Ureteroscopic extraction of calculus of ureter | | | | |
| | | OPCS4- M28·1 | Endoscopic laser fragmentation of calculus of ureter | | | | |
| | | OPCS4- M28·2 | Endoscopic fragmentation of calculus of ureter | | | | |
| | | OPCS4- M28·3 | Endoscopic extraction of calculus of ureter | | | | |
| | | OPCS4- M28·4 | Endoscopic catheter drainage of calculus of ureter | | | | |
| | | OPCS4- M28·8 | Other specified other endoscopic removal of calculus from ureter | | | | |
| | | OPCS4- M28·9 | Unspecified other endoscopic removal of calculus from ureter | | | | |
| | | OPCS4- M31·1 | Extracorporeal shock wave lithotripsy of calculus of ureter | | | | |
| | | OPCS4- M31·8 | Other specified extracorporeal fragmentation of calculus of ureter | | | | |
| | | OPCS4- M31·9 | Unspecified extracorporeal fragmentation of calculus of ureter | | | | |
| | | OPCS4- M26·1 | Nephroscopic laser fragmentation of calculus of ureter | | | | |
| | | OPCS4- M26·2 | Nephroscopic fragmentation of calculus of ureter NEC | | | | |
| | | OPCS4- M26·3 | Nephroscopic extraction of calculus of ureter | | | | |
| | | OPCS4- M28·5 | Endoscopic drainage of calculus of ureter by dilation of ureter | | | | |
| | | OPCS4- M28·8 | Other specified other endoscopic removal of calculus from ureter | | | | |

### Supplementary Table 3: Exclusion criteria for observational and genome-wide association analyses

| ICD-10 Codes | | ICD9 | OPCS 4 codes | | OPCS3 |
|---|---|---|---|---|---|
| E26·81 | Bartter syndrome | 255·13 | M39·1 | Open removal of calculus from bladder | 600·2 |
| E72·0 | Disorders of amino acid transport | 270 | M44·2 | Endoscopic extraction of calculus of bladder | |
| E21·0 | Hyperparathyroidism | 252 | M67·4 | Endoscopic removal of calculus from prostate | |
| E21·1 | Hyperparathyroidism | 252·02 | M75·8 | Open extraction of calculus from urethra | |
| E21·2 | Hyperparathyroidism | 252·08 | G27·1 | Gastric bypass surgery | 426·3 |
| E21·3 | Hyperparathyroidism | | G27·2 | Gastric bypass surgery | 426 |
| Q61·5 | Medullary sponge kidney | 753·17 | G27·3 | Gastric bypass surgery | |
| N25·8 | Type 1 renal tubular acidosis | | G27·4 | Gastric bypass surgery | |
| K50 | Inflammatory bowel disease | 558·9 | G27·5 | Gastric bypass surgery | |
| K51 | Inflammatory bowel disease | | G27·8 | Gastric bypass surgery | |
| K91·2 | Postsurgical malabsorption | 579·3 | G28·1 | Gastric bypass surgery | |
| Q62 | Congenital obstructive defects of the renal pelvis and malformations of the ureter | 753·29 | G28·2 | Gastric bypass surgery | |
| | | | G28·3 | Gastric bypass surgery | |
| E83·31 | Hereditary hypophosphatemic rickets with hypercalciuria and nephrolithiasis, osteoporosis, and hypophosphatemia | 275·3 | G28·4 | Gastric bypass surgery | |
| | | | G28·5 | Gastric bypass surgery | |
| E83·42 | Familial hypomagnesemia with hypercalciuria and nephrocalcinosis and Familial hypomagnesemia with hypercalciuria and nephrocalcinosis with ocular abnormalities | 275·2 | G28·8 | Gastric bypass surgery | |
| | | | G28·9 | Gastric bypass surgery | |
| E74·8 | Oxaluria and oxalosis | 271·4 | G31·1 | Gastric bypass surgery | |
| | | 271·8 | | | |
| N21·0 | Calculus in bladder | 594 | G31·2 | Gastric bypass surgery | |
| | | 594·1 | | | |
| N21·1 | Calculus in urethra | 594·2 | G31·3 | Gastric bypass surgery | |
| N21·8 | Other lower urinary tract calculus | 594·8 | G31·4 | Gastric bypass surgery | |
| N21·9 | Calculus of the lower urinary tract | 594·9 | G31·8 | Gastric bypass surgery | |
| | | | G31·9 | Gastric bypass surgery | |
| | | | G31·0 | Gastric bypass surgery | |
| | | | G32·1 | Gastric bypass surgery | |
| | | | G32·2 | Gastric bypass surgery | |
| | | | G32·3 | Gastric bypass surgery | |
| | | | G32·4 | Gastric bypass surgery | |
| | | | G32·8 | Gastric bypass surgery | |
| | | | G32·9 | Gastric bypass surgery | |
| | | | G32·0 | Gastric bypass surgery | |
| | | | G33·1 | Gastric bypass surgery | |
| | | | G33·2 | Gastric bypass surgery | |
| | | | G33·3 | Gastric bypass surgery | |
| | | | G33·6 | Gastric bypass surgery | |
| | | | G33·8 | Gastric bypass surgery | |
| | | | G33·9 | Gastric bypass surgery | |
| | | | G33·0 | Gastric bypass surgery | |

**Supplementary Table 4: Data sources for exposure and outcome variables in Mendelian randomization and association analyses**

| | Exposure variable | | Outcome variable | |
|---|---|---|---|---|
| Parameter | Source | Sample population | Source | Sample population |
| 2-hour glucose tolerance | Chen et al [1] | Meta-analysis of 24 of European ancestry | | |
| 24h urine calcium | | | Gary Curhan | NHSI/NHSII/HPFS, personal communication |
| 25-OH Vitamin D | Hannan et al | UK Biobank- European ancestry, personal communication | Hannan et al | UK Biobank- European ancestry, personal communication |
| Adjusted serum calcium* | Hannan et al | UK Biobank- European ancestry, personal communication | Hannan et al | UK Biobank- European ancestry, personal communication |
| APO-B | Richardson et al [2] | UK Biobank- European ancestry | | |
| BMD | Morris et al[3] | GWAS of 426,824 individuals in the UK Biobank- European ancestry | Morris et al[3] | GWAS of 426,824 individuals in the UK Biobank- European ancestry |
| BMI | Yengo et al [4] | Meta-analysis of UK Biobank GWAS and GIANT consortium studies. European ancestry | Yengo et al [4] | Meta-analysis of UK Biobank GWAS and GIANT consortium studies. European ancestry |
| CRP | Dehghan et al [5] | Meta-analysis of 11 studies, European ancestry | | |
| Fasting insulin | Chen et al [1] | Meta-analysis of 60 studies of European ancestry | | |
| Fasting glucose | Chen et al [1] | Meta-analysis of 71 studies including European ancestry | | |
| HbA1c | Chen et al [1] | Meta-analysis of 41 studies of European ancestry | | |
| HDL | Richardson et al [2] | UK Biobank- European ancestry | Richardson et al [2] | UK Biobank- European ancestry |
| HOMA-B | Dupuis et al [6] | Meta-analysis of 21 GWAS of European ancestry | | |
| HOMA-IR | Manning et al [7] | Joint meta-analysis of 29 studies using MAGIC consortium data, European ancestry, adjusted for BMI | | |
| Hypertension | Warren et al [8] | Meta-analysis of UK Biobank GWAS and exome data in 8 studies, European ancestry | | |
| KSD | GWAS presented in this manuscript | UK Biobank- European ancestry | GWAS presented in this manuscript | UK Biobank- European ancestry |
| LDL | Richardson et al [2] | UK Biobank- European ancestry | | |
| Serum phosphate | Hannan et al | UK Biobank- European ancestry, personal communication | Hannan et al | UK Biobank- European ancestry, personal communication |
| sIL-6R | Sun et al [9] | GWAS of INTERVAL dataset, European ancestry | | |
| T2D | Vijkovic et al [10] | Multi-ancestry meta-analysis of 228,499 cases and 1,178,783 controls in the Million Veteran Program (MVP), DIAMANTE, Biobank Japan, and other studies. European ancestry data used. | Vijkovic et al [10] | Multi-ancestry meta-analysis of 228,499 cases and 1,178,783 controls in the Million Veteran Program (MVP), DIAMANTE, Biobank Japan, and other studies. European ancestry data used. |
| Triglycerides | Richardson et al [2] | UK Biobank- European ancestry | | |
| Urate | Tin et al [11] | Meta-analysis of 288,649 participants of European ancestry | | |
| WC | Shungin et al[12] | GWAS meta-analyses of traits related to waist and hip circumferences in up to 224,459 individuals. European ancestry. | Shungin et al[12] | GWAS meta-analyses of traits related to waist and hip circumferences in up to 224,459 individuals. European ancestry. |
| WHR | Pulit et al [13] | Meta-analysis of UK Biobank GWAS and GIANT consortium studies. European ancestry | Pulit et al [13] | Meta-analysis of UK Biobank GWAS and GIANT consortium studies. European ancestry |

APO-B= apolipoprotein-B, BMD= bone mineral density, BMI= body mass index, CRP= C-reactive protein, GWAS= genome-wide association study, HDL= high-density lipoprotein, HOMA-B= homeostasis model assessment of β-cell function, HOMA-IR= homeostasis model assessment for insulin resistance, KSD= kidney stone disease, LDL= low-density lipoprotein, sIL-6R= serum IL-6 receptor, T2D= type 2 diabetes, WC= waist circumference, WHR= waist-to-hip ratio. *Total calcium + 0.0177x(46.3-albumin).


1. Chen J, Spracklen C, Marenne G, et al. The Trans-Ancestral Genomic Architecture of Glycaemic Traits. bioRxiv; 2020.
2. Richardson TG, Sanderson E, Palmer TM, et al. Evaluating the relationship between circulating lipoprotein lipids and apolipoproteins with risk of coronary heart disease: A multivariable Mendelian randomisation analysis. *PLoS medicine* 2020; **17**(3): e1003062.
3. Morris JA, Kemp JP, Youlten SE, et al. An atlas of genetic influences on osteoporosis in humans and mice. *Nature genetics* 2019; **51**(2): 258-66.
4. Yengo L, Sidorenko J, Kemper KE, et al. Meta-analysis of genome-wide association studies for height and body mass index in approximately 700000 individuals of European ancestry. *Human molecular genetics* 2018; **27**(20): 3641-9.
5. Dehghan A, Dupuis J, Barbalic M, et al. Meta-analysis of genome-wide association studies in >80 000 subjects identifies multiple loci for C-reactive protein levels. *Circulation* 2011; **123**(7): 731-8.
6. Dupuis J, Langenberg C, Prokopenko I, et al. New genetic loci implicated in fasting glucose homeostasis and their impact on type 2 diabetes risk. *Nature genetics* 2010; **42**(2): 105-16.

7. Manning AK, Hivert MF, Scott RA, et al. A genome-wide approach accounting for body mass index identifies genetic variants influencing fasting glycemic traits and insulin resistance. *Nature genetics* 2012; **44**(6): 659-69.
8. Warren HR, Evangelou E, Cabrera CP, et al. Genome-wide association analysis identifies novel blood pressure loci and offers biological insights into cardiovascular risk. *Nature genetics* 2017; **49**(3): 403-15.
9. Sun BB, Maranville JC, Peters JE, et al. Genomic atlas of the human plasma proteome. *Nature* 2018; **558**(7708): 73-9.
10. Vujkovic M, Keaton JM, Lynch JA, et al. Discovery of 318 new risk loci for type 2 diabetes and related vascular outcomes among 1.4 million participants in a multi-ancestry meta-analysis. *Nature genetics* 2020; **52**(7): 680-91.
11. Tin A, Marten J, Halperin Kuhns VL, et al. Target genes, variants, tissues and transcriptional pathways influencing human serum urate levels. *Nature genetics* 2019; **51**(10): 1459-74.
12. Shungin D, Winkler TW, Croteau-Chonka DC, et al. New genetic loci link adipose and insulin biology to body fat distribution. *Nature* 2015; **518**(7538): 187-96.
13. Pulit SL, Stoneman C, Morris AP, et al. Meta-analysis of genome-wide association studies for body fat distribution in 694 649 individuals of European ancestry. *Human molecular genetics* 2019; **28**(1): 166-74.

**Supplementary Table 5: UK Biobank Study Population for genome-wide association studies.**

| Cohort | Number of Samples | Female (%) | Male (%) |
|---|---|---|---|
| Kidney stone | 8,504 | 2,871 (33·8) | 5,633 (66·2) |
| Control | 388,819 | 212,081 (54·5) | 176,738 (45·5) |

**Supplementary Table 6: Single nucleotide polymorphisms significantly associated with kidney stone disease from male sex genome-wide association study**

| Chromosome | Position[a] | rsID | Annotation | EA | Non-Effect Allele | EAF[b] | Info | OR (95% CI) | P | Candidate Gene |
|---|---|---|---|---|---|---|---|---|---|---|
| 1 | 21826530 | rs115239632 | Intergenic | T | C | 0·05 | 1 | 1·44 (1·31-1·59) | $2{\cdot}7\times10^{-13}$ | *ALPL* |
| 1 | 21840129 | rs869179 | Intronic | G | A | 0·67 | 1 | 1·12 (1·08-1·17) | $1{\cdot}3\times10^{-8}$ | *ALPL* |
| 1 | 21893344 | rs1256332 | Intronic | A | C | 0·19 | 1 | 1·20 (1·14-1·26) | $2{\cdot}0\times10^{-12}$ | *ALPL* |
| 5 | 176798040 | rs56235845 | Intronic | G | T | 0·36 | 0·99 | 1·18 (1·13-1·22) | $9{\cdot}5\times10^{-16}$ | *SLC34A1* |
| 6 | 160611103 | rs78693187 | Intronic | C | T | 0·04 | 1 | 1·32 (1·22-1·53) | $2{\cdot}1\times10^{-8}$ | *SLC22A2* |
| 7 | 30957616 | rs2299905 | Intronic | T | A | 0·31 | 0·99 | 1·12 (1·08-1·17) | $4{\cdot}9\times10^{-8}$ | *AQP1* |
| 7 | 142605221 | rs4252512 | Intergenic | C | T | 0·02 | 0·99 | 1·51 (1·31-1·74) | $1{\cdot}2\times10^{-8}$ | *TRPV5* |
| 13 | 42758805 | rs9590676 | Intronic | C | T | 0·40 | 1 | 1·12 (1·08-1·17) | $1{\cdot}9\times10^{-9}$ | *DGKH* |
| 15 | 53997089 | rs578595 | Intronic | C | A | 0·49 | 1 | 1·11 (1·07-1·16) | $2{\cdot}3\times10^{-8}$ | *WDR72* |
| 16 | 20392332 | rs77924615 | Intronic | A | G | 0·22 | 0·98 | 1·15 (1·10-1·21) | $3{\cdot}2\times10^{-9}$ | *UMOD* |
| 20 | 52742680 | rs111707488 | Intergenic | A | AGT | 0·32 | 0·94 | 1·13 (1·09-1·19) | $3{\cdot}2\times10^{-9}$ | *CYP24A1* |
| 21 | 37818871 | rs2776288 | Intronic (ncRNA) | A | G | 0·66 | 0·99 | 1·15 (1·11-1·20) | $7{\cdot}5\times10^{-13}$ | *CLDN14* |
| 22 | 23410918 | rs13054904 | Intronic | A | T | 0·30 | 1 | 1·17 (1·12-1·22) | $4{\cdot}4\times10^{-13}$ | *BCR* |

[a]Based on NCBI Genome Build 37 (hg19).[b]The effect allele frequency in participants with KSD. Three independent signals were identified at the *ALP* locus. SNP= single nucleotide polymorphism

**Supplementary Table 7: Single nucleotide polymorphisms significantly associated with kidney stone disease from female sex genome-wide association study**

| Chromosome | Positiona | rsID | Annotation | EA | Non-Effect Allele | EAFb | Info | OR (95% CI) | P | Candidate Gene |
|---|---|---|---|---|---|---|---|---|---|---|
| 7 | 27617940 | 7:27617940_AT_A | Intronic | AT | A | 0·32 | 0·95 | 1·18 (1·12-1·26) | $1{\cdot}5{\times}10^{-8}$ | *HIBADH* |
| 20 | 52731402 | rs6127099 | Intergenic | A | T | 0·75 | 0·99 | 1·19 (1·12-1·26) | $1{\cdot}4{\times}10^{-9}$ | *CYP24A1* |
| 21 | 37818871 | rs2776288 | Intronic (ncRNA) | A | G | 0·66 | 0·99 | 1·17 (1·12-1·24) | $2{\cdot}7{\times}10^{-9}$ | *CLDN14* |

[a]Based on NCBI Genome Build 37 (hg19).[b]The effect allele frequency in participants with KSD. SNP= single nucleotide polymorphism

### Supplementary Table 8: Mendelian randomization analyses of effects of the metabolic syndrome on kidney stone disease

| Exposure | Outcome | Inverse-variance weighted | | | Intercept | | MR-Egger | | Weighted median | | Contamination mixture | |
|---|---|---|---|---|---|---|---|---|---|---|---|---|
| | | OR (95% CI) | p | p-adjusted | Beta (95% CI) | p | OR (95% CI) | p | OR (95% CI) | p | OR (95% CI) | p |
| **Serum LDL concentration** | KSD | **0·96 (0·86-1·06)** | **0·37** | **0·56** | 0·00 (0·00-0·00) | 0·25 | 0·89 (0·77-1·04) | 0·15 | - | - | - | - |
| **Serum TG concentration** | KSD | 1·14 (1·04-1·25) | 3·9×10$^{-3}$ | 0·01 | **0·00 (0·00-0·00)** | **6·1×10$^{-3}$** | **0·98 (0·85-1·13)** | **0·78** | - | - | - | - |
| **Serum HDL concentration** | KSD | 1·03 (0·95-1·13) | 0·46 | 0·62 | **0·00 (0·00-0·00)** | **6·4×10$^{-3}$** | **1·19 (1·04-1·37)** | **9·9×10$^{-3}$** | - | - | - | - |
| **Serum HDL adjusted for serum calcium concentration*** (mvMR) | KSD | 1·01 (0·92-1·12) | 0·82 | 0·85 | **0·00 (0·00-0·00)** | **0·04** | **1·02 (0·92-1·12)** | **0·76** | - | - | - | - |
| **Serum calcium* adjusted for serum HDL concentration** (mvMR) | KSD | 1·78 (1·56-2·03) | 3·6×10$^{-18}$ | 1·1×10$^{-16}$ | | | **2·01 (1·69-2·38)** | **1·5×10$^{-15}$** | - | - | - | - |
| **T2D** | KSD | 1·12 (1·07-1·17) | 2·8×10$^{-6}$ | 1·5×10$^{-5}$ | **0·00 (0·00-0·00)** | **0·01** | **1·00 (0·91-1·10)** | **0·99** | 1·06 (1·00-1·13) | 0·06 | 1·07 (1·00-1·15) | 0·06 |
| **Fasting glucose adjusted for BMI** | KSD | **1·00 (0·82-1·21)** | **0·98** | **0·98** | 0·00 (0·0-0·00) | 0·95 | 0·99 (0·71-1·38) | 0·94 | - | - | - | - |
| **Fasting insulin adjusted for BMI** | KSD | **1·32 (0·74-2·35)** | **0·36** | **0·56** | 0·00 (-0·02-0·01) | 0·44 | 2·54 (0·43-14·90) | 0·30 | - | - | - | - |
| **HbA1c** | KSD | **1·26 (0·96-1·66)** | **0·10** | **0·21** | 0·00 (0·00-0·00) | 0·88 | 1·30 (0·82-2·05) | 0·27 | - | - | - | - |
| **HOMA-IR** | KSD | **0·90 (0·35-2·35)** | **0·84** | **0·85** | 0·04 (-0·01-0·08) | 0·11 | 0·02 (0·00-2·40) | 0·11 | - | - | - | - |
| **HOMA-B** | KSD | **1·20 (0·74-1·93)** | **0·46** | **0·62** | -0·01 (-0·03-0·01) | 0·26 | 2·69 (0·61-11·83) | 0·91 | - | - | - | - |
| **2-hour glucose tolerance adj for BMI** | KSD | **0·92 (0·70-1·22)** | **0·57** | **0·71** | 0·02 (0·00-0·04) | 1·32 | 0·54 (0·26-1·14) | 0·11 | - | - | - | - |
| **BMI adjusted for WHR and T2D** (mvMR) | KSD | **1·10 (0·96-1·27)** | **0·19** | **0·36** | 0·00 (0·00-0·00) | 0·34 | 1·17 (0·97-1·43) | 0·11 | - | - | - | - |
| **WHR adjusted for BMI and T2D** (mvMR) | KSD | **1·64 (1·35-2·00)** | **5·0×10$^{-7}$** | **3·1×10$^{-6}$** | | | 1·65 (1·36-2·01) | 1·3×10$^{-8}$ | - | - | - | - |
| **T2Dadjusted for BMI and WHR** (mvMR) | KSD | **0·98 (0·94-1·02)** | **0·33** | **0·52** | | | 0·98 (0·93-1·02) | 0·29 | - | - | - | - |
| **Systolic blood pressure** | KSD | **1·01 (0·98-1·04)** | **0·41** | **0·58** | 0·00 (-0·02-0·02) | 0·91 | 1·02 (0·89-1·17) | 0·78 | - | - | - | - |
| **Diastolic blood pressure** | KSD | **1·01 (0·97-1·04)** | **0·64** | **0·71** | 0·00 (-0·01-0·01) | 0·84 | 0·99 (0·86-1·15) | 0·93 | - | - | - | - |
| **Pulse pressure** | KSD | **1·01 (0·99-1·04)** | **0·39** | **0·58** | 0·00 (-0·01-0·01) | 0·84 | 1·00 (0·92-1·10) | 0·97 | - | - | - | - |
| | | **Beta (95% CI)** | **p** | **p-adjusted** | **Beta (95% CI)** | **p** | **Beta (95% CI)** | **p** | | | | |
| **Serum HDL concentration** | Serum calcium concentration* | **0·04 (0·02-0·07)** | **5·5×10$^{-5}$** | **2·4×10$^{-4}$** | 0·00 (0·00-0·00) | 0·09 | 0·07 (0·03-0·10) | 1·3×10$^{-4}$ | - | - | - | - |
| **Serum HDL concentration** | Serum 25-OH vitamin D concentration | 0·01 (-0·02-0·03) | 0·63 | 0·71 | **0·00 (0·00-0·00)** | **1·5×10$^{-7}$** | **-0·06 (-0·10--0·03)** | **1·9×10$^{-4}$** | - | - | - | - |
| **WHR** | Serum HDL concentration | **-0·39 (-0·47--0·32)** | **6·9×10$^{-25}$** | **4·3×10$^{-23}$** | 0·00 (0·00-0·00) | 0·71 | -0·36 (-0·57--0·14) | 1·3×10$^{-3}$ | - | - | - | - |

*Albumin adjusted serum calcium concentration; 25-OH vitamin D = hydroxyvitamin D; adj=adjusted BMI = body mass index; CI=confidence interval; HDL = high density lipoprotein; KSD = kidney stone disease; LDL = low density lipoprotein; OR=odds ratio for outcome per 1 standard deviation increase in genetically-instrumented exposure variable; p-adjusted = p value adjusted for multiple testing using false discovery rate method; WHR = waist-to-hip ratio; T2D= type 2 diabetes; TG = triglyceride; mvMR=multivariable Mendelian randomization.
Bold text highlights the sensitivity analysis to be interpreted after considering the estimate of the intercept.

**Supplementary Table 9: Mendelian randomization analyses of effects of inflammation-related phenotypes on kidney stone disease**

| Exposure | Outcome | Inverse-variance weighted | | | Intercept | | MR-Egger | |
|---|---|---|---|---|---|---|---|---|
| | | OR (95% CI) | p | p-adjusted | Beta (95% CI) | p | OR (95% CI) | p |
| CRP | KSD | **1·11 (0·93-1·32)** | **0·25** | 0·44 | 0·00 (-0·02-0·01) | 0·70 | 1·17 (0·84-1·64) | 0·35 |
| sIL-6R | KSD | **1·01 (0·99-1·03)** | **0·31** | 0·51 | 0·00 (-0·01-0·00) | 0·39 | 1·02 (0·99-1·06) | 0·21 |
| APOB | KSD | **0·97 (0·89-1·06)** | **0·56** | 0·71 | 0·00 (0·00-1·00) | 0·60 | 0·95 (0·84-1·08) | 0·44 |

APO-B= apolipoprotein-B; CI=confidence interval.; CRP= C-reactive protein; KSD = kidney stone disease; OR=odds ratio for outcome per 1 standard deviation increase in genetically-instrumented exposure variable; p-adjusted = p value adjusted for multiple testing using false discovery rate method; sIL-6R= serum IL-6 receptor

Bold text highlights the sensitivity analysis to be interpreted after considering the estimate of the intercept.

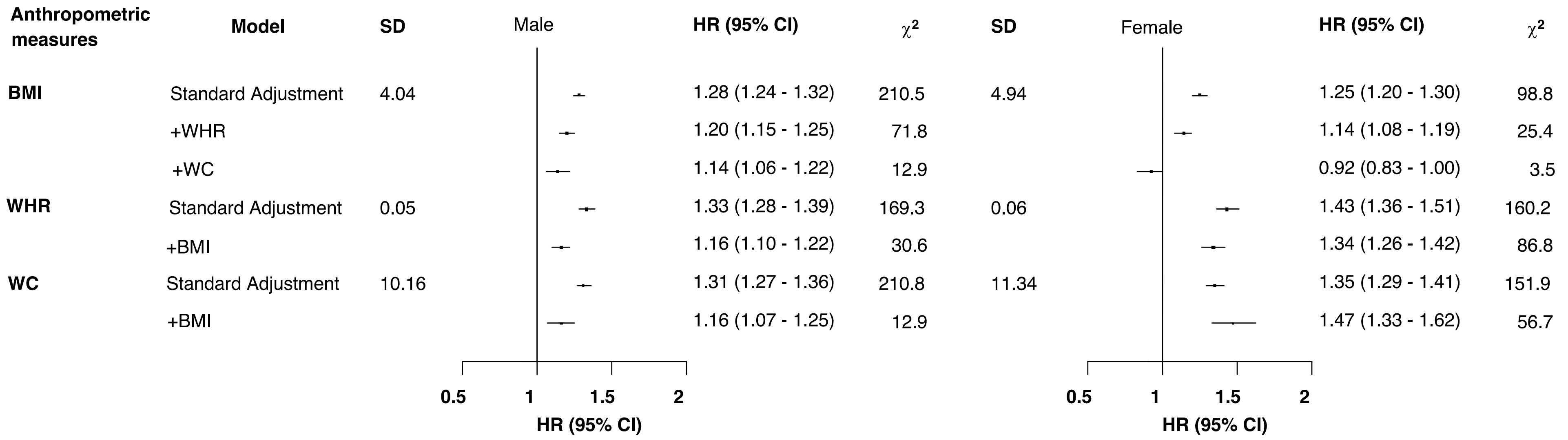


**Supplementary Figure 1: Hazard ratio (HR) for incident kidney stone disease (KSD) and 95% confidence interval (CI) per standard deviation (SD) change in body mass index (BMI), waist-to-hip ratio (WHR) and waist circumference (WC)**

HR per SD stratified by age-at-risk, and ethnicity, and adjusted for Townsend Deprivation Index, smoking and alcohol drinking (standard adjustments), with further adjustments for other anthropometric measures where indicated. Analyses exclude participants with pre-existing KSD (or conditions known to predispose to KSD) at baseline, and those with missing or outlying values in anthropometric variables or key covariates, leaving 478,405 participants. The variance of the category-specific log risk determines the CI.

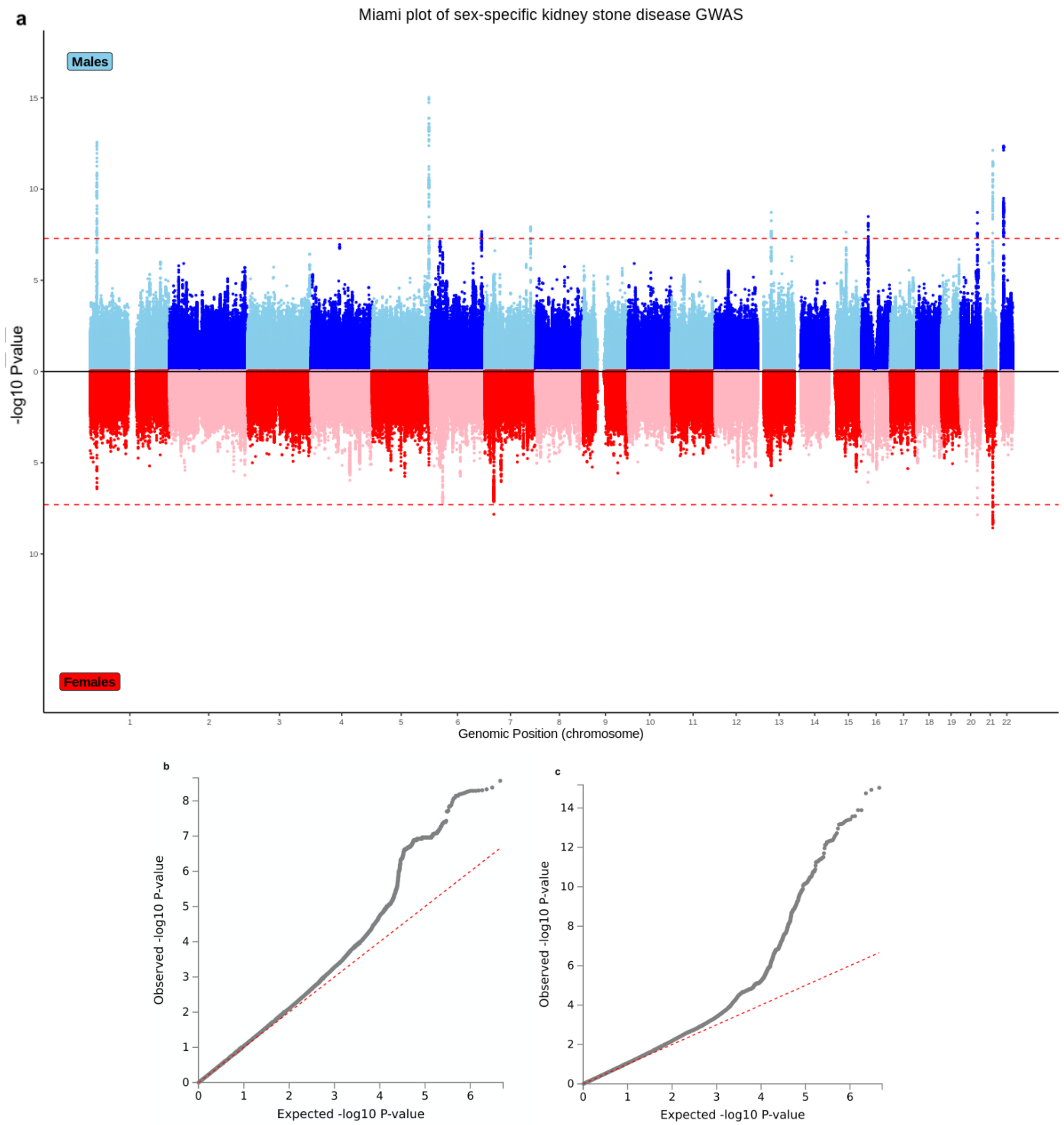


**Supplementary Figure 2: Results of sex-specific genome wide association study in kidney stone disease**

a. Miami plot illustrating the results of sex-specific genome-wide association studies (GWAS) in kidney stone disease (KSD) in the UK Biobank. Analysis of genetically male individuals included 5,633 cases and 176,738 controls, blue. Analysis of genetically female individuals included 2,871 cases and 212,081 controls, red. Genome-wide p values (-log10) are plotted against their respective positions on each of the autosomes. The horizontal red line shows the genome-wide significance threshold of 5·0 × 10−8.

b. Quantile-quantile plot of observed vs. expected p-values for female-specific GWAS of KSD. λGC= 1·05, LD score regression (LDSC) intercept=1·003, attenuation ratio =0·283

c. Quantile-quantile plot of observed vs. expected p-values for male-specific GWAS of KSD. λGC= 1·05, LD score regression (LDSC) intercept=1·005, attenuation ratio =0·147

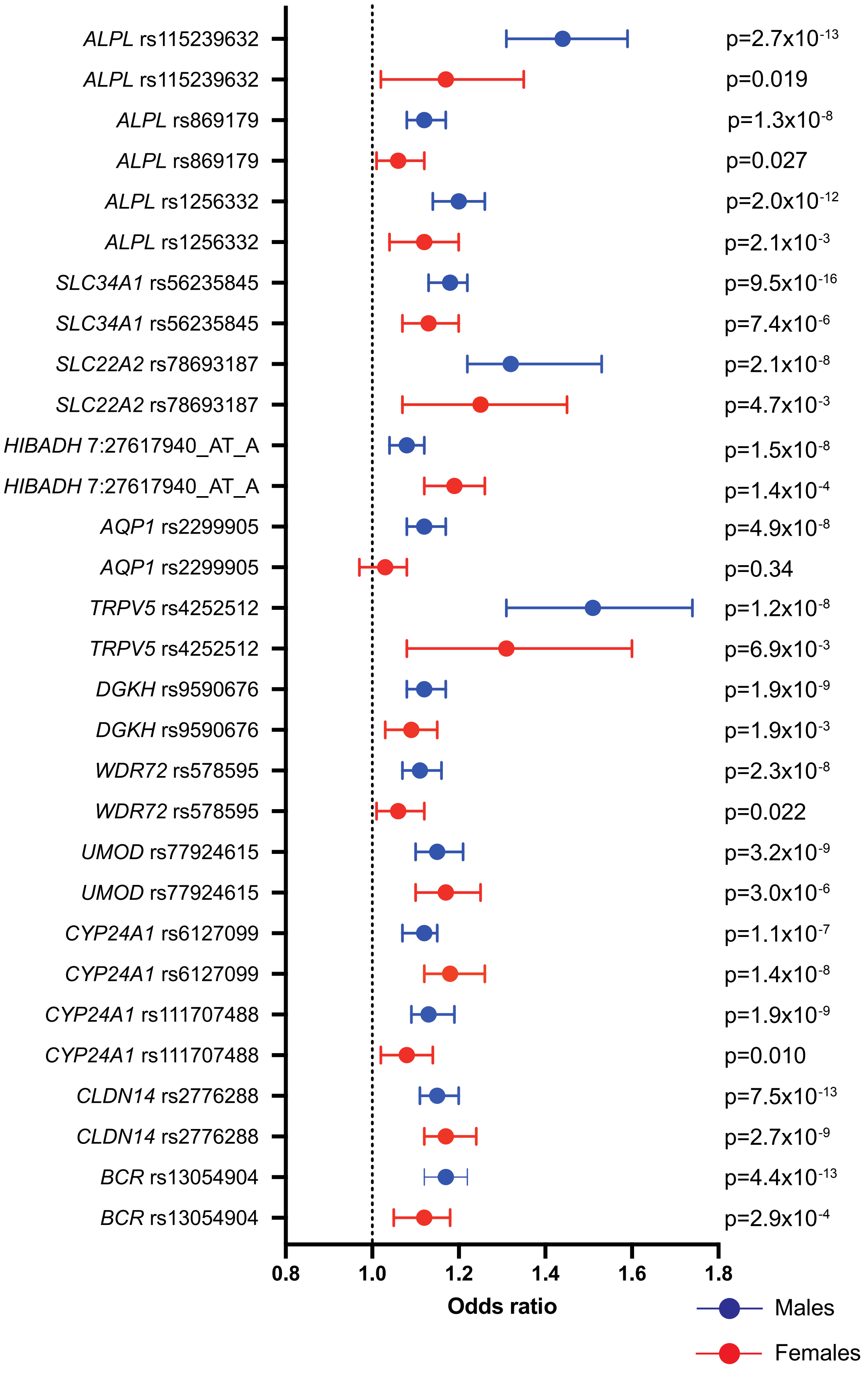


**Supplementary Figure 3: Sex-specific odds ratios, 95% confidence intervals and p values for significant hits in genome wide association studies in males or females.**
Male data is represented in blue, female data is represented in red.

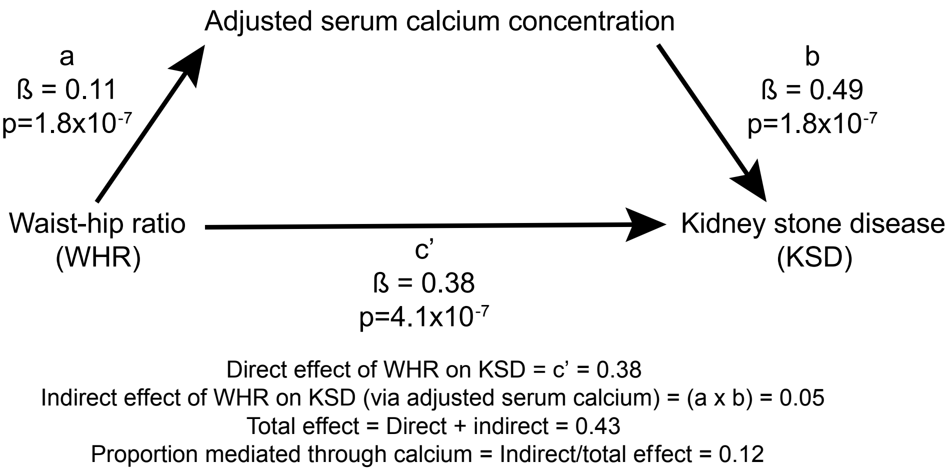


**Supplementary Figure 4: Effects of waist-to-hip ratio on serum calcium concentration and kidney stone disease.**
Mediation Mendelian randomisation. ß= regression coefficient for each MR analysis, p= p value adjusted for multiple testing using false discovery rate method

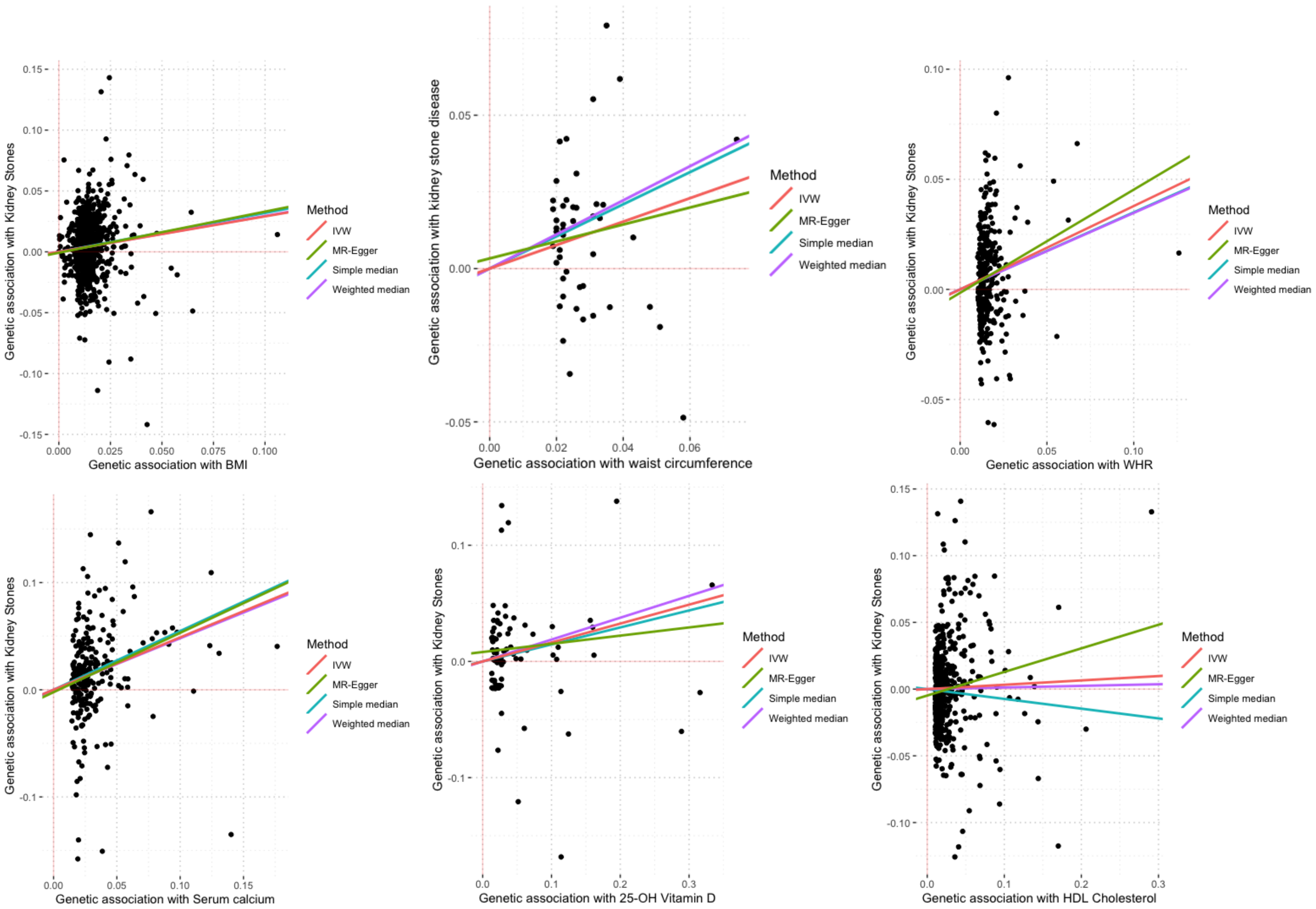
Genetic association with Kidney Stones
Genetic association with BMI
Method
IVW
MR-Egger
Simple median
Weighted median
Genetic association with kidney stone disease
Genetic association with waist circumference
Method
IVW
MR-Egger
Simple median
Weighted median
Genetic association with Kidney Stones
Genetic association with WHR
Method
IVW
MR-Egger
Simple median
Weighted median
Genetic association with Kidney Stones
Genetic association with Serum calcium
Method
IVW
MR-Egger
Simple median
Weighted median
Genetic association with Kidney Stones
Genetic association with 25-OH Vitamin D
Method
IVW
MR-Egger
Simple median
Weighted median
Genetic association with Kidney Stones
Genetic association with HDL Cholesterol
Method
IVW
MR-Egger
Simple median
Weighted median

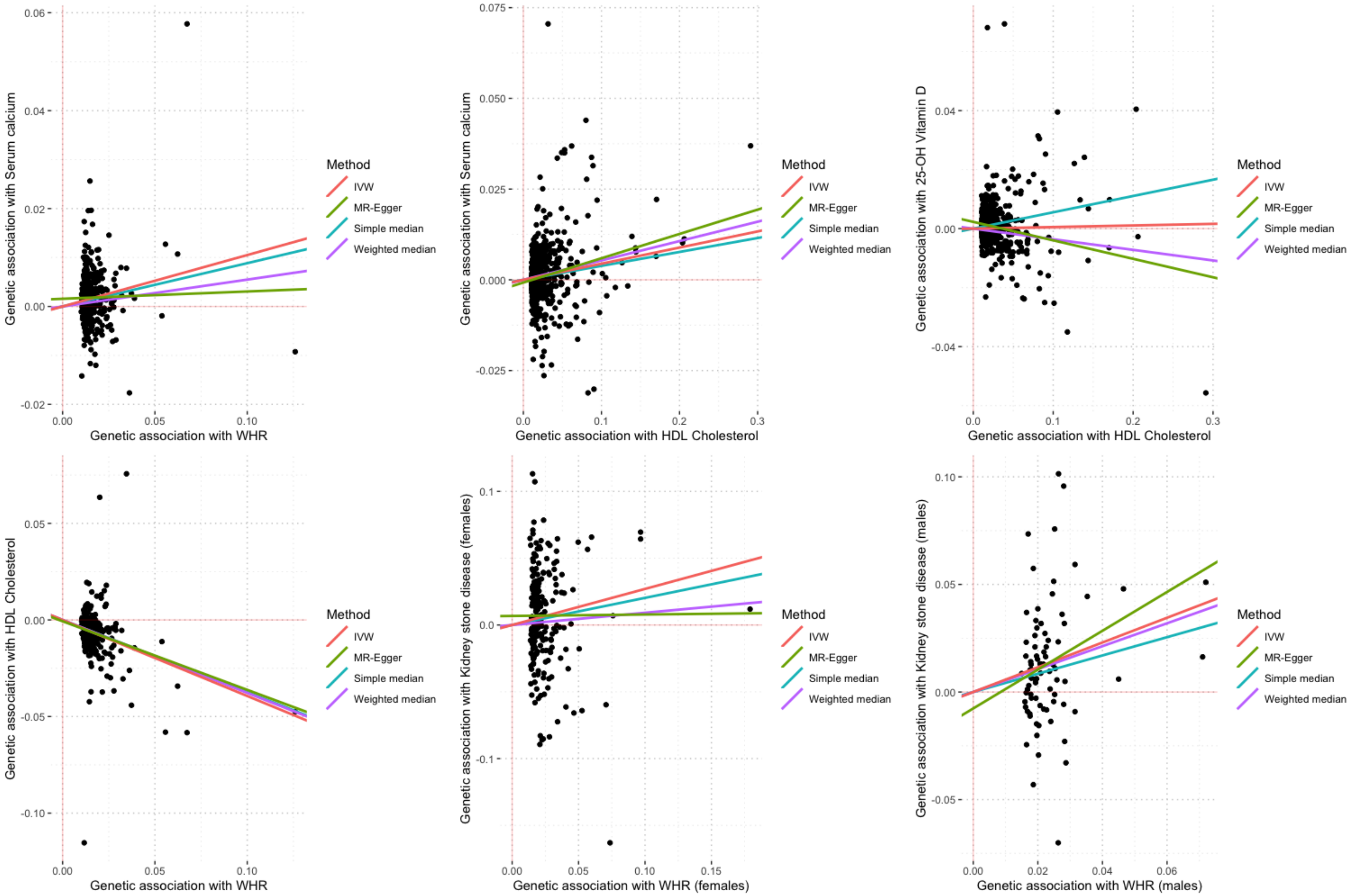
Genetic association with Serum calcium
Genetic association with WHR
Genetic association with Serum calcium
Genetic association with HDL Cholesterol
Genetic association with 25-OH Vitamin D
Genetic association with HDL Cholesterol
Genetic association with HDL Cholesterol
Genetic association with WHR
Genetic association with Kidney stone disease (females)
Genetic association with WHR (females)
Genetic association with Kidney stone disease (males)
Genetic association with WHR (males)
Method
IVW
MR-Egger
Simple median
Weighted median

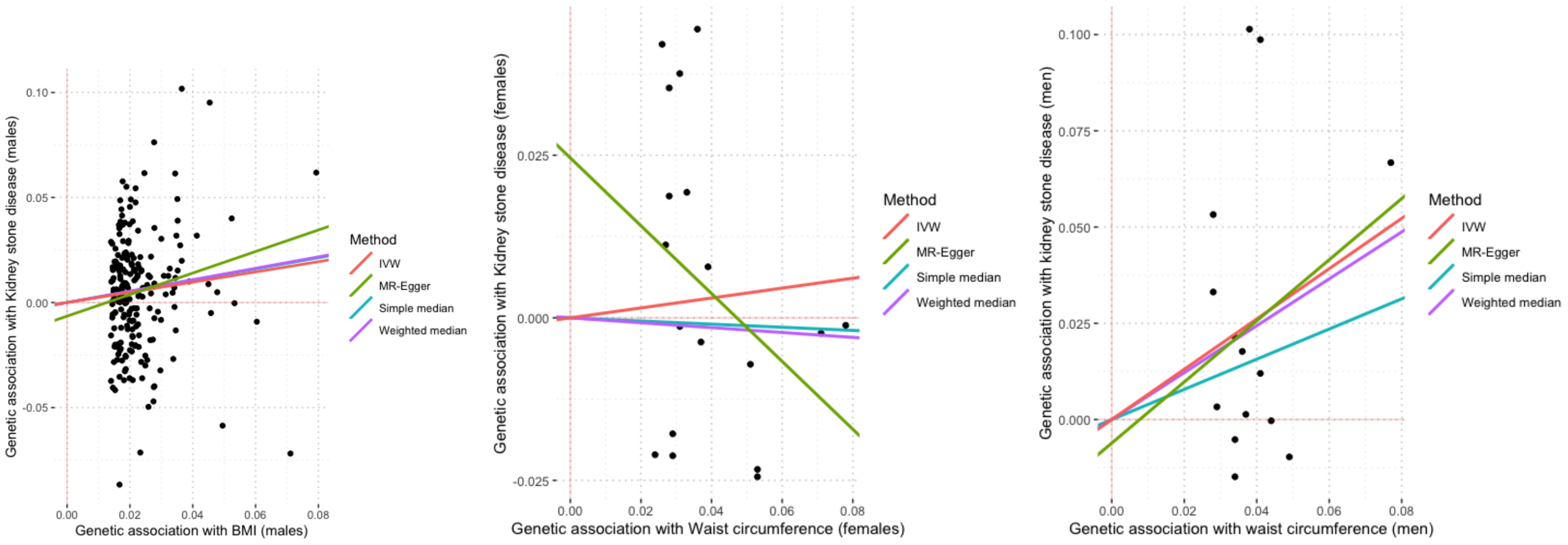


**Supplementary Figure 5: Significant relationships identified on Mendelian randomization**

Scatter plots of significant anthropometric, biochemical, and metabolic exposure variables verses kidney stone disease in the UK Biobank. Regression estimates are shown by coloured lines rescribed in each figure legend.

BMI= body mass index, HDL= high density lipoproteins, IVW= inverse-variance weighted, WHR= waist-to-hip ratio
